# Disorder-Free Data are All You Need: Inverse Supervised Learning for Broad-Spectrum Head Disorder Detection

**DOI:** 10.1101/2023.10.10.23296794

**Authors:** Yuwei He, Yuchen Guo, Jinhao Lyu, Liangdi Ma, Haotian Tan, Wei Zhang, Guiguang Ding, Hengrui Liang, Jianxing He, Xin Lou, Qionghai Dai, Feng Xu

## Abstract

**BACKGROUND:** The development of artificial intelligence (AI)-based medical systems heavily relies on the collection and annotation of sufficient data containing disorders. However, the preparation of data with complete disorder types and adequate annotations presents a significant challenge, limiting the diagnostic capabilities of existing AI-based medical systems. This study introduces a novel AI-based system that accurately detects a broad spectrum of disorders without requiring any disorder-containing data.

**METHODS:** We obtained a training dataset of 21,429 disorder-free head computed tomography (CT) scans and proposed a learning algorithm called Inverse Supervised Learning (ISL). This algorithm learns and understands disorder-free samples instead of disorder-contained ones, enabling the identification of all types of disorders. We also developed a diagnosis and visualization software for clinical usage based on the system’s ability to provide visually understandable clues.

**RESULTS:** The system achieved Area Under the Curve (AUC) values of 0.883, 0.868, and 0.866 on retrospective (127 disorder types, 9,967 scans), prospective (117 disorder types, 3,054 scans), and cross-center (46 disorder types, 554 scans) datasets, respectively. These results demonstrate that the system can detect far more disorder types than previous AI-based systems. Furthermore, the ISL-based systems achieved AUC values of 0.893 and 0.895 on pulmonary CT and retinal optical coherence tomography (OCT), respectively, demonstrating that ISL can generalize well to non-head and non-CT images.

**CONCLUSIONS:** Our novel AI-based system, utilizing ISL, can accurately and broadly detect disorders without requiring disorder-containing data. This system not only outperforms previous AI-based systems in terms of disorder detection but also provides visually understandable clues, enhancing its clinical utility. The successful application of ISL to non-head and non-CT images further demonstrates its potential for broad-spectrum medical applications. (Funded by National Key R&D Program of China, National Natural Science Foundation of China)

---

Over the past decade, artificial intelligence (AI) has made significant strides and has been applied in various fields. In the medical field, the accumulation of medical image data has enabled many AI diagnostic techniques to achieve radiologist-level performance in recognizing, classifying, and quantifying specific diseases. For example, AI has been used for cerebral hemorrhage recognition^1^ and COVID-19 recognition^2^ from CT images. These breakthroughs have led us to envision that AI diagnostic techniques can assist in clinical decision-making from medical images and alleviate the severe shortage of expert radiologists in many areas and hospitals.

Despite the significant progress made in AI techniques, there is still a gap between these techniques and real clinical decision-making. Current AI techniques primarily focus on recognizing specific types of disorders from input medical data. However, for a clinical decision-making workflow, the most basic and essential task is to identify all possible disorder types that could be diagnosed from the medical image. For instance, in the case of brain CT, more than one hundred types of disorders could be diagnosed from it. Therefore, a decision-making diagnostic system for brain CT must be capable of detecting a broad spectrum of disorders, as missing the detection of any disorder type is unacceptable. Existing medical AI techniques are developed based on widely-used AI paradigms, which involve deciding the disorder types to be handled, collecting sufficient disorder-contained samples, and constructing recognition/localization/segmentation models for the disorders. This paradigm works well when the disorder types are limited, and the samples are easily accessible. However, developing a broad-spectrum disorder detection system using this paradigm requires collecting data and constructing models for all types of disorders, which is extremely difficult and inefficient, especially for unusual diseases. Therefore, it is impractical touse the widely-used AI paradigms to achieve real clinical decision-making.

To address the challenges mentioned above, we propose a novel AI solution called Inverse Supervised Learning (ISL). Instead of using disorder-contained data, which requires hundreds of disorder types and a large number of samples for each type, we use disorder-free medical images for supervision. In theory, we use the opposite problem (detecting no-disorder samples) to replace the original problem (detecting all types of disorders). Therefore, instead of training hundreds of models to recognize all possible types of disorders, we train just one model to understand the concept of disorder-free fully. Consequently, all disorders can be identified as they differ from the disorder-free samples used in training. With our paradigm, the challenges mentioned above are fully resolved as there is no need for samples of all possible disorder types.

To achieve ISL, we utilize the traditional computer vision task of image inpainting in a novel framework. Image inpainting aims to restore the content of a partially missing image based on the context of non-missing information. Specifically, in this case, an image inpainting network is trained to replenish masked regions in a medical image, where the replenished content always reflects healthy tissue because the training dataset contains only disorder-free medical images. If any disorder exists in the image and the disorder region is masked off, the reconstructed disorder-free image would be significantly different from the original one. Conversely, for an image without disorders, no matter which region is masked, the reconstructed image should always be similar to the original one as they are both healthy and consistent with the rest of the healthy images. By masking, inpainting, and comparing all the image regions, ISL can detect various types of disorders and locate the disorder regions. Our proposed solution, ISL, (1) does not require the deliberate collection of data for any disorder type; (2) ensures that the data used to develop systems are easily accessible; (3) does not require experts to manually annotate the data; (4) enables the developed system to recognize broad-spectrum disorders rather than specific ones; and (5) provides experts with clinical clues, such as disorder locations.

In this study, we utilized ISL to construct a system for broad-spectrum disorder detection on unenhanced brain computed tomography (CT) scans.^3, 4^ CT is a first-line diagnostic modality for assessing brain abnormalities due to its quick acquisition and non-invasive nature. The ISL-based system was developed using only disorder-free head CT images. It achieved expert-level accuracies on a retrospective dataset with 127 disorder types and a prospective dataset with 116 disorder types, surpassing the number of detectable disorder types in previous works. We also applied ISL to build two additional systems: one for pulmonary disorder detection in CT images and another for retinal disorder detection in optical coherence tomography (OCT) images. The results demonstrate that ISL can generalize well to non-brain and non-CT-based disorder detection.

## Results

### Building an ISL based system for clinically applicable broad-spectrum head disorder detection

Our proposed ISL-based disorder detection system for brain CT comprises a de-disorder network (DeDN), a disorder recognition network (DRN), and a disorder locating module. Firstly, a CT image is processed with specific window width and window locations and then fed into the DeDN to generate its corresponding de-disorder image. Next, we obtain a difference image by subtracting the original and generated images. Finally, the difference image is inputted into the DRN to determine whether any disorder exists in the image. Additionally, the disorder locating module can be used to locate the disorder. Our goal is to provide an effective tool that can assist physicians and researchers in quickly identifying images that may contain disorders from a large volume of CT images for further analysis and diagnosis.

To develop the system, we collected CT scans from the Chinese PLA General Hospital (PLAGH), a leading national hospital that serves patients throughout China. We constructed a training dataset of 21,429 healthy brain CT scans (March 2012 - July 2019) retrieved from the picture archiving and communication systems (PACS). The retrieval process involved matching the fixed description (“No abnormality is observed”) of healthy CT scans with historical diagnosis reports, resulting in a training dataset that was efficiently obtained without requiring expert effort or disorder annotation.

### Performance on the broad-spectrum head disorder detection

To evaluate the system, we obtained a retrospective test dataset from the PLAGH (9,967 scans, 88.23% with 127 types of disorders, March 2012 - July 2019) and a prospective test dataset from the PLAGH (3,054 scans, 88.70% with 116 types of disorders, July 2019 - August 2021). To demonstrate the clinical applicability of our system, we counted all types of disorders described in clinical reports from the PLAGH using a rule-based NLP algorithm and manual selection by radiologists. We sorted out 127 and 116 types of disorders for testing, respectively. To our knowledge, these test datasets have the broadest coverage of head disorder types. The number of scans for each disorder is shown in Supplementary Table 1 and 2.

We employed a disorder-contained/free classification testing strategy for each type of disorder, with testing carried out at the scan-level. This means that the system predicted whether the entire scan contained any disorder or not. Scan-level classification is practical for clinical use as it enables radiologists to quickly identify the presence of disorders, which is particularly useful in emergency treatment.^5^ The label of each scan was determined using disorder-related keywords in its associated report and then confirmed by radiologists based on the report and CTimages.

For the retrospective and prospective test datasets, the area under the receiver operating characteristic curve (AUC) with 95% confidence interval (CI) for the two datasets, along with the true positive rate (TPR), false positive rate (FPR), and the overall receiver operating characteristic (ROC) curves, are presented in Supplementary Table 1, 2, 4, and Supplementary Figure 1. Additionally, Supplementary Table 1 and 2 also present the sensitivity and specificity with 95% CI for the disorders. On the retrospective dataset, the system achieved an AUC *>* 0.95 for 43 disorders and an AUC *>* 0.90 for 74 disorders. On the prospective dataset, the system achieved an AUC *>* 0.95 for 30 disorders and an AUC *>* 0.90 for 50 disorders. These results demonstrate that our system is capable of detecting a broad spectrum of disorders in brain CT.

### Analysis of lesion detection efficacy by size and urgency of treatment

In our comprehensive analysis, we delved into the challenges of disorder detection. We identified two primary categories of challenging cases in disorder detection: those that are small and easily missed, and those that do not require immediate treatment, which may also be overlooked due to their subtler characteristics. To conduct a thorough analysis, we divided the cases into three groups based on these dimensions.

In terms of lesion size, we classified the cases as large, medium, or small. The classification outcomes are detailed in Supplementary Table 7. We computed the Area Under the Curve (AUC) values with 95% CI (Table 1) and plotted ROC curves (Figure 1) for each size category. The AUC results for different lesion sizes demonstrate AUC accuracies of 0.941, 0.943, and 0.887 for large, medium, and small lesions, respectively. These figures underscore our model’s high accuracy in detecting even smaller lesions, maintaining a commendable level of recognition precision.

**Table 1.** Performance for disorder types across different lesion sizes.

| Performance vs. Lesion sizes |  |  |  |
| --- | --- | --- | --- |
|  | AUC | Sensitivity | Specificity |
| Large | 0.941 (0.941, 0.942) | 0.883 (0.882, 0.885) | 0.865 (0.864, 0.867) |
| Medium | 0.943 (0.943, 0.944) | 0.885 (0.883, 0.886) | 0.875 (0.873, 0.876) |
| Small | 0.887 (0.887, 0.888) | 0.885 (0.884, 0.887) | 0.771 (0.770, 0.773) |

**Figure 1.**
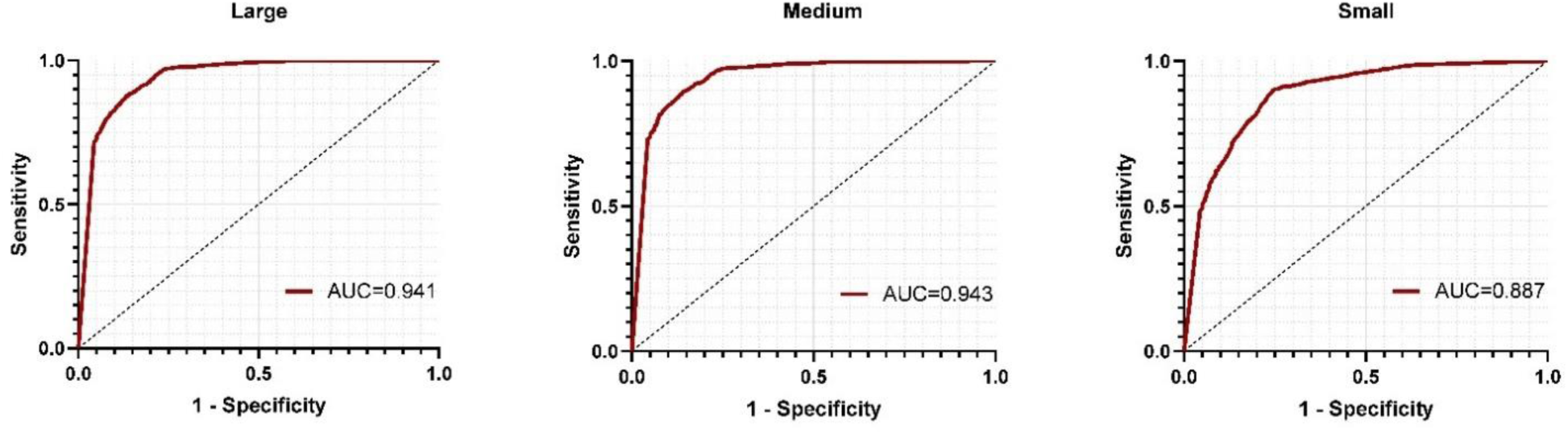
ROC curves for disorder types across different lesion sizes.

When classifying based on the urgency of treatment, we sorted the cases into high, medium, and low urgency levels, calculating the corresponding AUC values for each. The categories were defined as follows: *Emergency intervention*. This group encompasses severe disorders necessitating immediate medical attention, such as certain cancers and other conditions that could be life-threatening. *Selective intervention*. Disorders in this category may not require urgent treatment but could necessitate medical intervention as they evolve. *Non-intervention*. This group includes disorders that generally do not require treatment and have minimal impact on patient quality of life. Detailed classification results are shown in Supplementary Table 8.

The AUC accuracies and ROC curves for lesions of high, medium, and low urgency are shown in Table 2 and Figure 2. The AUC results were 0.946, 0.859, and 0.861, respectively. These results indicate that our model is proficient in identifying lesions with varying degrees of urgency, effectively recognizing even those with less pronounced features.

**Table 2.** Performance for disorder types based on urgency of treatment.

| Performance vs. Urgency of treatment |  |  |  |
| --- | --- | --- | --- |
|  | AUC | Sensitivity | Specificity |
| High | 0.942 (0.941, 0.942) | 0.859 (0.858, 0.861) | 0.897 (0.895, 0.898) |
| Medium | 0.853 (0.853, 0.854) | 0.849 (0.848, 0.851) | 0.727 (0.726, 0.729) |
| Low | 0.859 (0.859, 0.860) | 0.838 (0.836, 0.840) | 0.737 (0.736, 0.739) |

**Figure 2.**
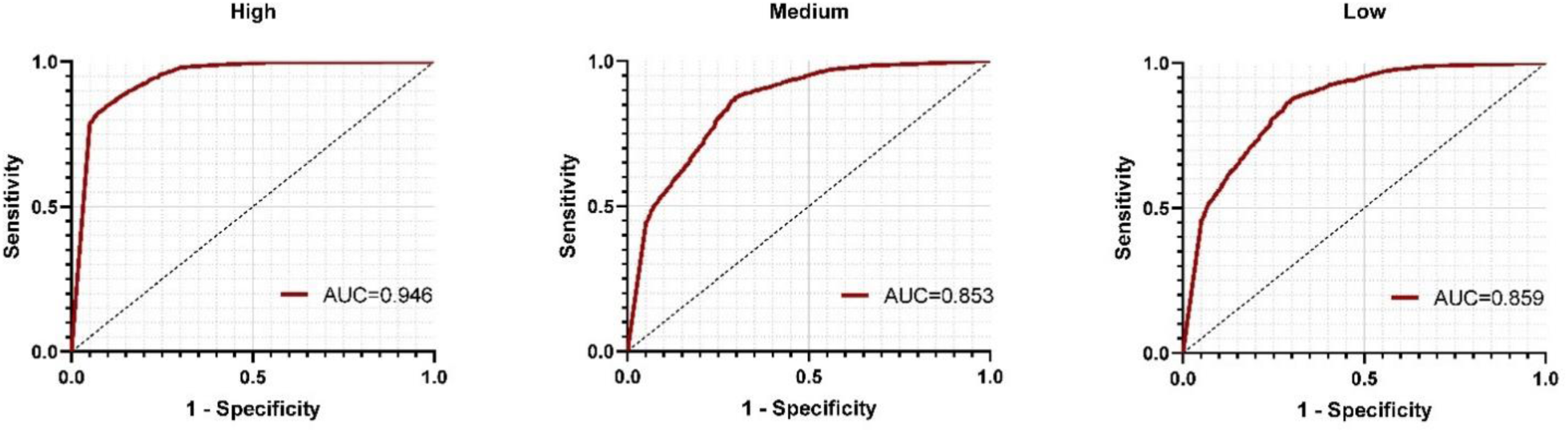
ROC curves for disorder types based on urgency of treatment.

### Evaluation of system generalizability

To be practical, an AI-based system should be able to generalize to new data from different centers and hospitals. In order to evaluate the generalizability of the ISL-based system, we constructed a cross-center test dataset from the Brain Hospital of Hunan Province (BHHP), which served as an independent test cohort from PLAGH. This dataset consisted of 554 scans, of which 59.01% had 46 different types of disorders. It is worth noting that in the cross-center experiment, we made efforts to collect as much available data as possible to ensure the comprehensiveness of the tested disorders. However, this approach resulted in a smaller number of samples for certain disorders (e.g., the total sample size for Basal Ganglia Cerebral Infarction was 5). As a result, the performance of these specific disorder types may deviate when compared to the retrospective test set.

The AUCs with 95% CI for the 46 types of disorders, along with the overall ROC curve are presented in Supplementary Table 3 and Supplementary Figure 1. The average AUC was 0.866, which was only 0.017 lower than that of the retrospective intra-center test. These results demonstrate the generalizability of the system across different centers.

### Evaluation of improving expert performance

In clinical practice, a computer-aided diagnosis (CAD) system should provide understandable clues to support prediction results. Our model can quickly and intuitively locate the disorder region based on the generated de-disorder image, as shown in Figure 3a. Additionally, we developed a CAD software for clinical use, as shown in Figure 3b. The software takes a CT scan as input and outputs possible disorder regions, improving the diagnosis performance of radiologists.

**Figure 3.**
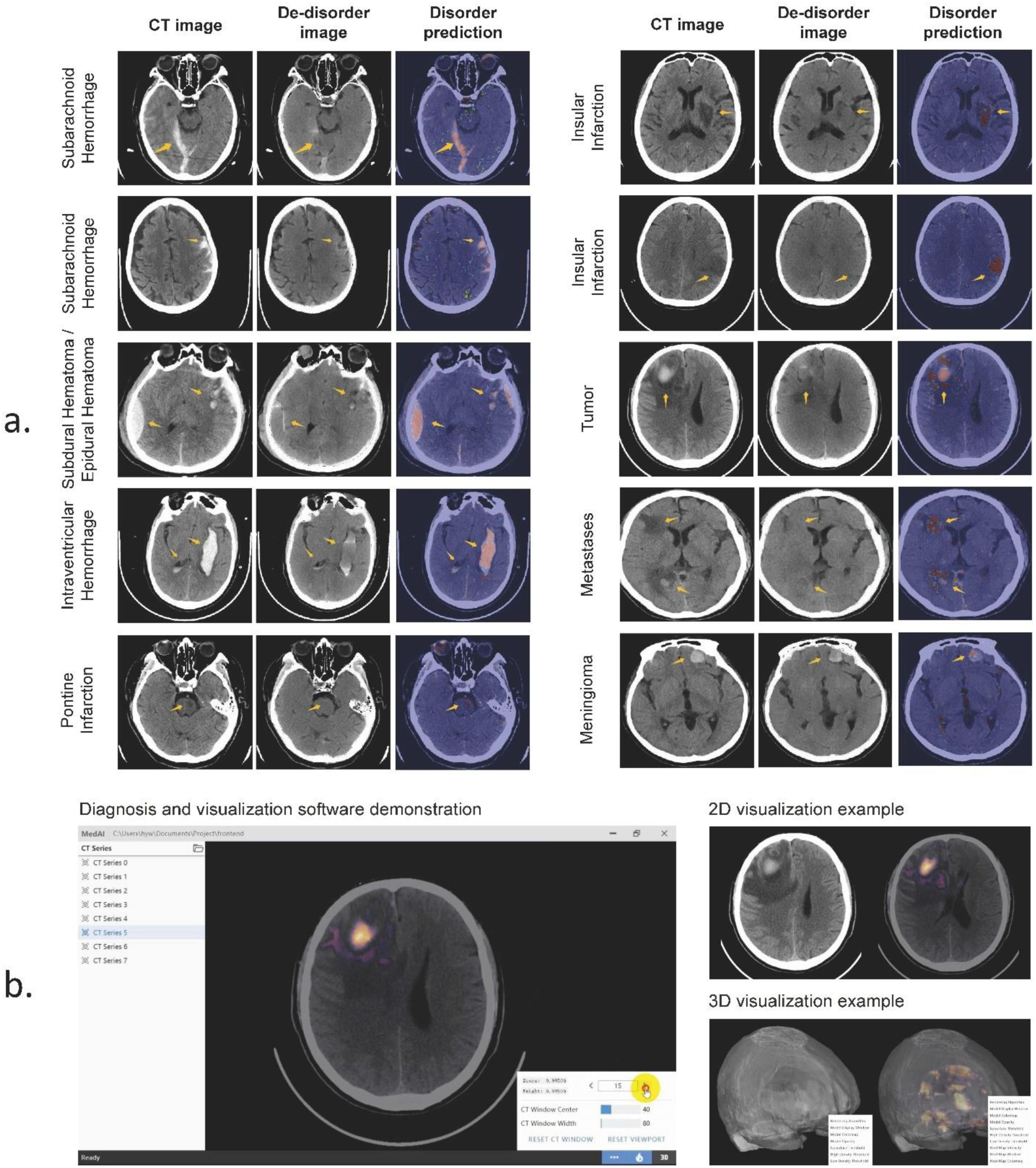
Visualization examples for head disorder detection. **a)** Typical examples of our system’s performance are shown, including the original CT image, the corresponding de-disorder image generated by our system, and the heatmap indicating the probability of containing disorders. Warmer colors in the heatmap indicate a higher probability of disorders. The heatmaps provide visual clues to the system’s decision. **b)** We also developed a diagnosis and visualization software that takes a CT scan as input and outputs possible disorder locations in the form of a heatmap. The heatmap can be displayed on a 2D slice or on a 3D reconstruction scan.

To quantitatively evaluate the improvement, we conducted an experiment involving four radiologists with diverse levels of experience, ranging from 5 to 14 years. Each radiologist was tasked with independently reviewing a set of 300 randomly chosen samples from our cross-center test dataset, which comprised 100 cases with identified disorders and 200 cases deemed healthy. Initially, the radiologists performed their assessments without the support of our software, relying solely on their expertise. Subsequently, we introduced the diagnostic suggestions provided by our software to examine its influence on the radiologists’ ability to diagnose accurately.

The incorporation of software’s insights led to a notable enhancement in diagnostic precision. The average sensitivity across the four radiologists increased by 0.035, while the specificity saw a marginal improvement of 0.006. the advancements are visually represented in Figure 4.

**Figure 4.**
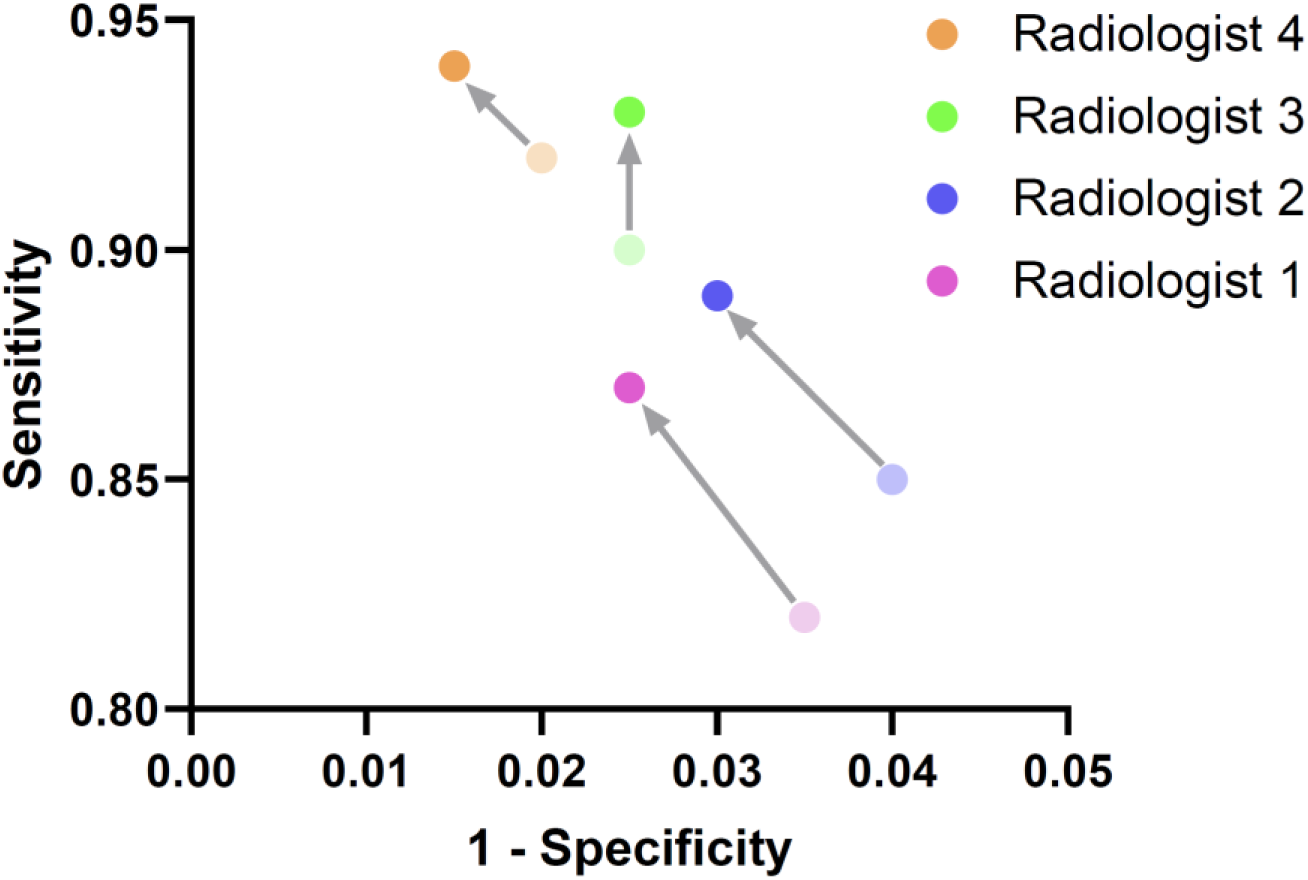
The performance of four radiologists before and after considering the system recommendation. The radiologists have 5 (pink), 7 (blue), 10 (green) and 14 (orange) years of working experience.

The observed improvements underscore the potential of our software to serve as a valuable tool for radiologists, particularly in the accurate detection of disorders. The integration of our software into the diagnostic workflow promises to refine disorder screening processes and support radiologists in delivering more precise and reliable diagnoses. The radiologists reported that the system effectively reduced their workload by accurately identifying a broad spectrum of disorders, and contributed to lowering the rate of missed diagnoses. They appreciated the system’s ability to provide visually understandable clues, which greatly assisted in their diagnosis process.

### Analysis of system explainability

Our system not only detects the disorder location in a slice but also provides the disorder distribution in a scan. Figure 5b shows two example scans with different disorder distributions. Based on the distribution curves supplied by our system, we can observe that the disorders in the two scans have centralized and dispersive distributions,respectively. Figure 5c shows the average disorder distributions for some typical disorders in the retrospective dataset. The average distributions are close to the occurrence frequency at different brain tissues in reality, demonstrating the effectiveness of the system’s explainability.

**Figure 5.**
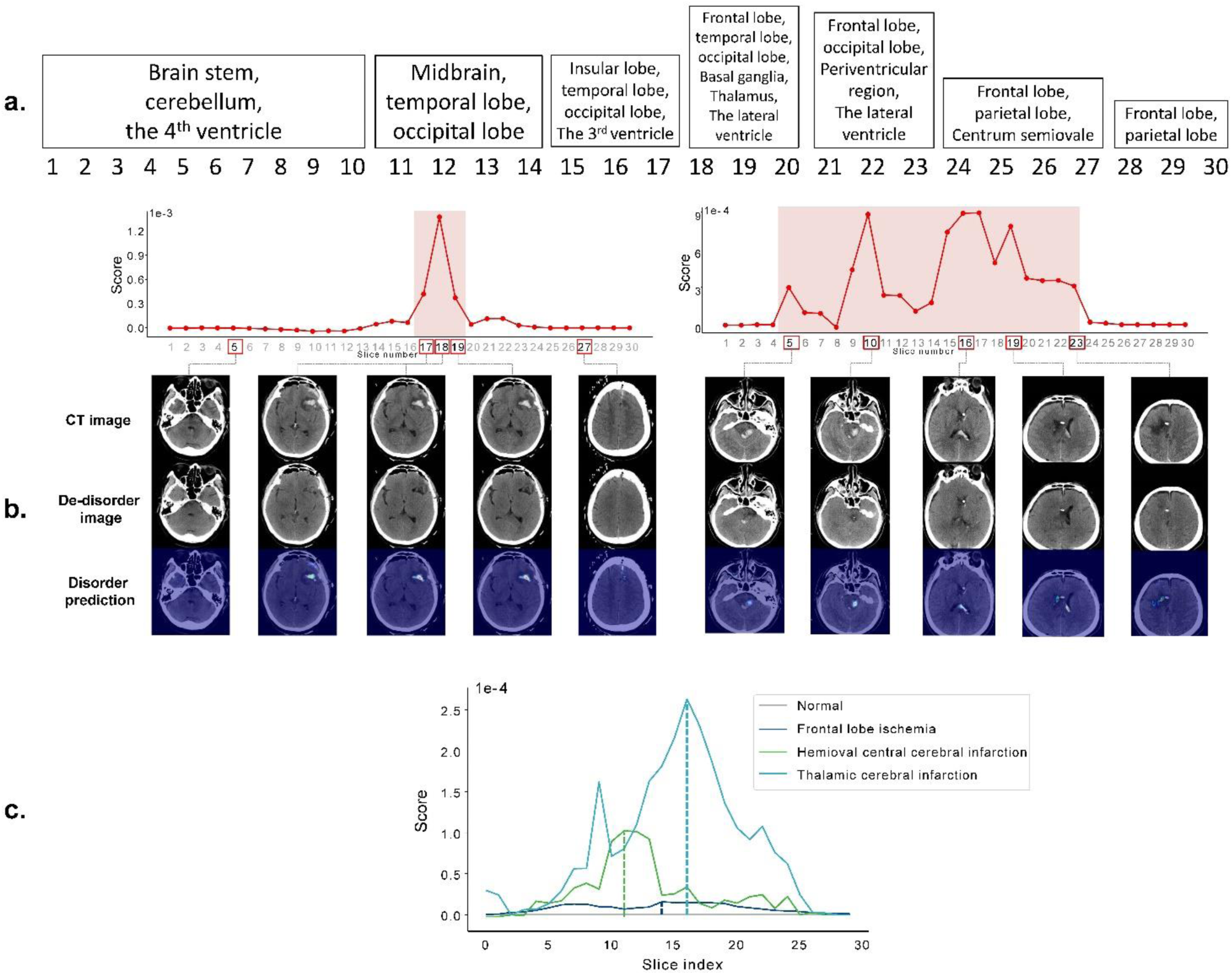
Patterns of disorder distribution in CT scans. The x-axis of the graph represents the slice index of a CT scan, while the y-axis represents a slice’s abnormal score. A higher score indicates a greater likelihood of lesions in the slice. **a)** The correspondence between slice indexes and brain tissues is shown. **b)** Two example scans with dispersive and centralized distributions are presented. **c)** The average disorder distributions of some typical disorders in the retrospective dataset are displayed.

### Performance contributions from different modules

This section elaborates on the reasons for adopting each module and demonstrates their contributions to the final performance. The results are presented in Figure 6.

**Figure 6.**
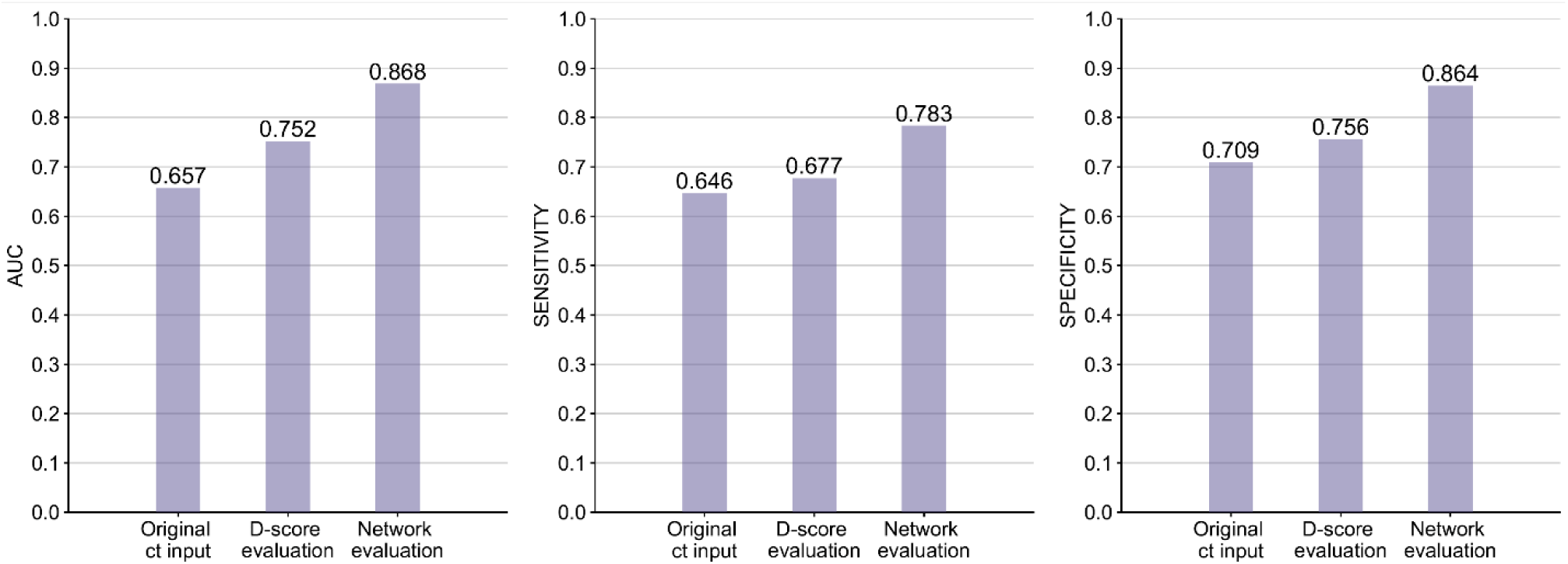
Iterative performance improvement by de-disorder network (DeDN) and disorder recognition network (DRN). **Original ct input**: original CT slices + DRN; **D-score evaluation**: DeDN + pixel value sum; **Network evaluation**: DeDN + DRN. The comparison was performed on the prospective test dataset.

#### Performance contribution from de-disorder network

In ISL, the evaluation of the probability of disorder containment depends on the difference image **x***_dif_*, where **x***_dd_* is a de-disorder image generated by a de-disorder network (DeDN). Original medical images are too complex for a system to learn disorder-related information solely based on them. Therefore, we do not directly apply original images for evaluation. Using difference images for evaluation is more intuitive, as the greater the difference between the original and de-disorder images, the higher the probability of disorder containment.

To numerically demonstrate the effectiveness of the difference image generated by the DeDN, we also applied the original-image-based method for evaluation. As shown in Figure 6, on the prospective test dataset, the average AUC with 95% confidence interval is 0.657 and 0.752, respectively, where the result from the original-image-based method (**Original CT input**) is significantly lower than that from the difference-image-based method (**D-score evaluation**). The improved result highlights the value of the DeDN.

#### Performance contribution from disorder recognition network

After obtaining the difference image **x***_dif_* with a DeDN, we used a disorder recognition network (DRN) to evaluate the probability of disorder containment. Although we could determine the probability directly based on the pixel value sum of the difference image, we did not adopt this strategy. This is because a DeDN cannot produce perfectly healthy tissue, meaning that even for a healthy area, the pixel value sum of that area may still be positive. As a result, the accumulated pixel value sum of all healthy areas would negatively influence the probability evaluation.

Instead, we used the pixel sum-based evaluation (**D-score evaluation**) and DRN-based evaluation (**Network evaluation**) based on the difference image, as shown in Figure 6. The average AUC of the two methods on the prospective dataset were 0.752 and 0.868, respectively, demonstrating the superiority of the DRN.

### Evaluation of inverse-supervised learning generalizability

To assess the generalizability of ISL across different body parts and medical image types, we employed it to develop two additional systems. The first system is designed for detecting pulmonary disorders in CT images, while the second system is designed for detecting retinal disorders in optical coherence tomography (OCT) images.

#### Performance of pulmonary disorder detection

In addition to brain CT, we developed an ISL-based system for detecting disorders in pulmonary CT scans. The data used for system development were collected from the First Affiliate Hospital of Guangzhou Medical University (FAHGMU), another leading national hospital that serves patients from across China. We constructed a training dataset consisting of 3,410 healthy pulmonary CT scans and a test dataset that included 6 types of pulmonary disorders (82 pneumothorax, 86 pneumonia, 96 bronchiectasis, 88 bullae, 82 atelectasis, and 46 effusion), as well as 600 healthy scans. The AUCs and detection examples for each type of disorder are presented in Table 3 and Figure 7a. The average AUC was 0.893, indicating that ISL can generalize well across different body parts.

**Table 3.**
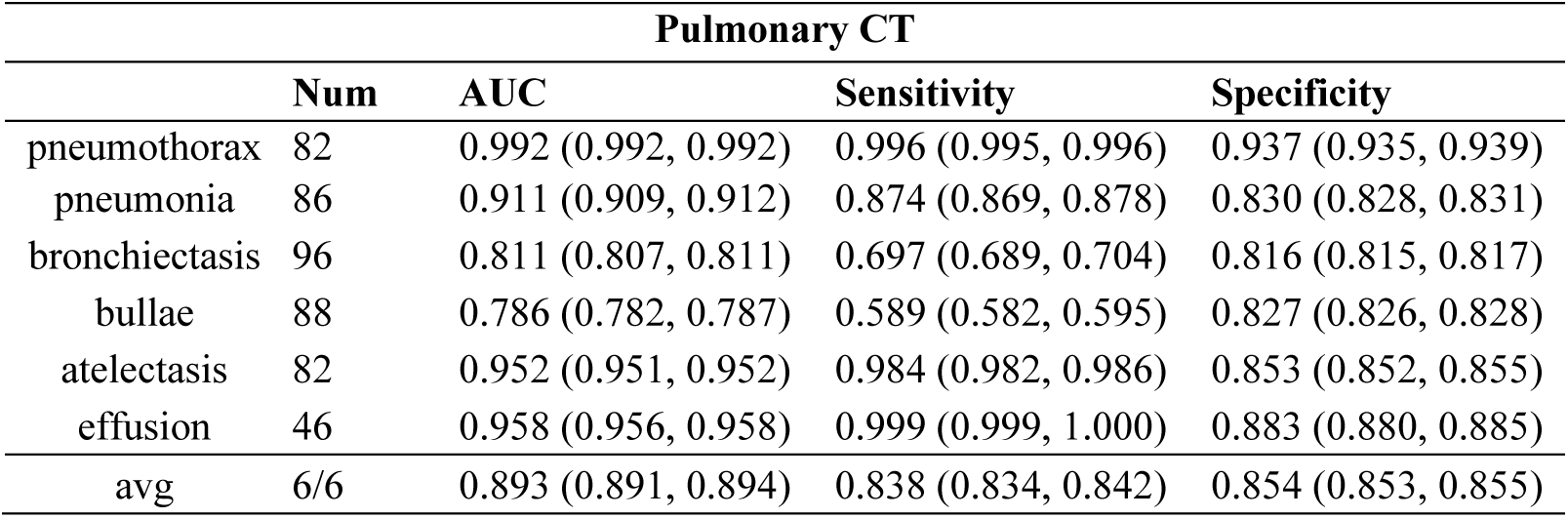
Performance on the pulmonary CT test dataset for pulmonary disorder detection.

**Figure 7.**
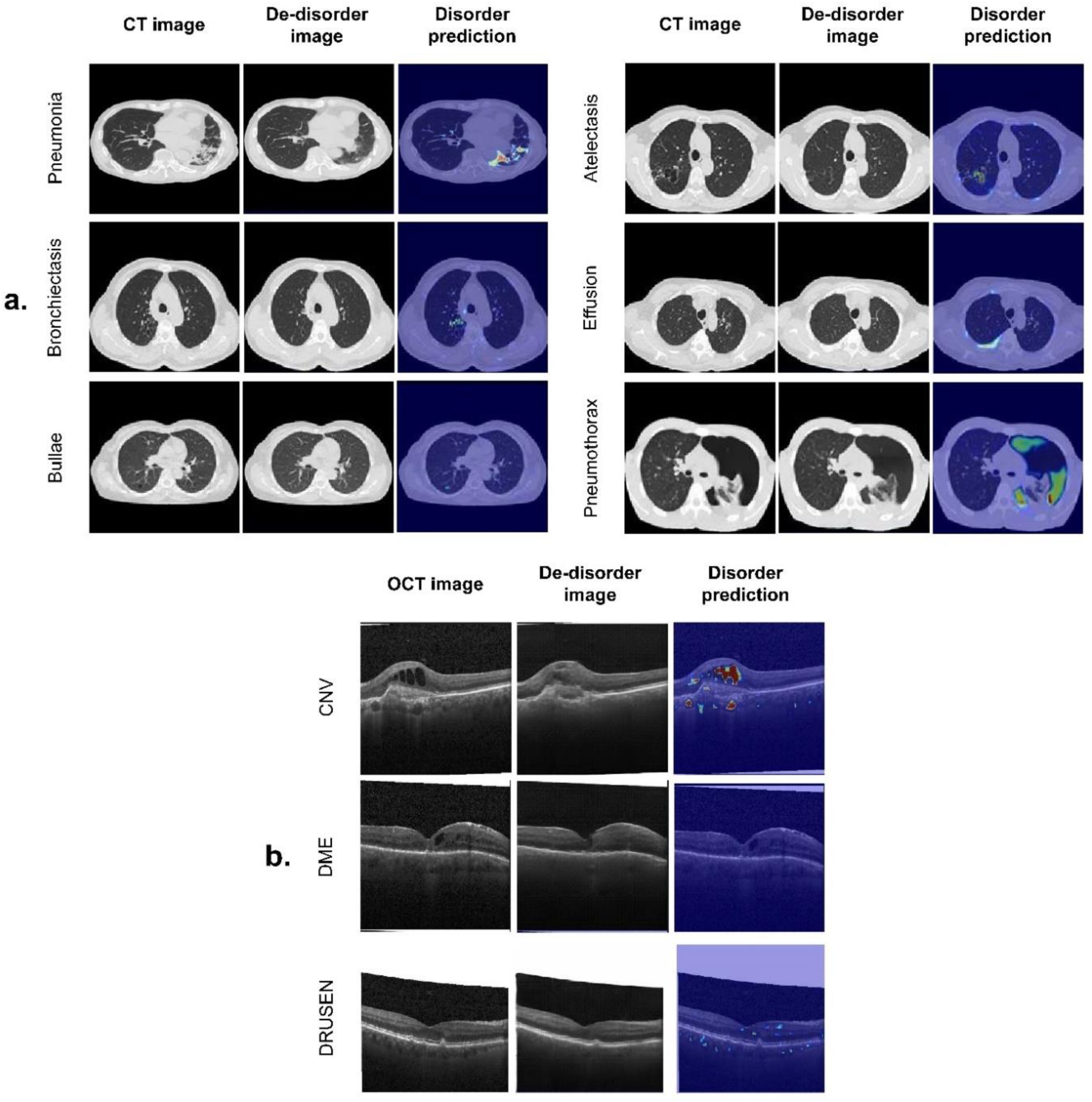
Examples of our system on the CT-based pulmonary disorder detection and the OCT-based retinal disorder detection.

#### Performance of retinal disorder detection

To demonstrate the ability of ISL to generalize across different medical image types, we developed a retinal disorder detection system based on optical coherence tomography (OCT) images. We used the dataset collected by Kermany et al.,^6^ which includes 108,312 images (37,206 with choroidal neovascularization, 11,349 with diabetic macular edema, 8,617 with drusen, and 51,140 normal). Following the development procedure of ISL, we used only the normal OCT images as the training dataset. The model was tested with 1,000 images (250 from each category) from 633 patients, as in Kermany et al.^6^ The AUCs with 95% confidence interval (CI) on the scan-level are summarized in Table 4. The AUCs for choroidal neovascularization (CNV), diabetic macular edema (DME), and drusen were 0.939, 0.913, and 0.827, respectively. Despite being developed using only normal OCT images, our system achieved clinically acceptable results, indicating that ISL is applicable to different medical image types. Detection examples for each type of disorder are shown in Figure 7b.

**Table 4.** Performance on the retinal OCT test dataset for retinal disorder detection.

| Retinal OCT |  |  |  |  |
| --- | --- | --- | --- | --- |
|  | Num | AUC | Sensitivity | Specificity |
| CNV | 1,220 | 0.939 (0.938, 0.940) | 0.847 (0.844, 0.851) | 0.904 (0.900, 0.907) |
| DME | 1,600 | 0.913 (0.912, 0.914) | 0.838 (0.835, 0.841) | 0.890 (0.888, 0.893) |
| DRUSEN | 1,220 | 0.827 (0.826, 0.829) | 0.760 (0.757, 0.764) | 0.762 (0.758, 0.765) |
| avg | 3/3 | 0.895 (0.894, 0.896) | 0.817 (0.814, 0.820) | 0.855 (0.852, 0.858) |

## Discussion

We introduced a learning strategy called inverse supervised learning (ISL) and utilized it to develop a head disorder detection system that requires no disorder data or annotation during the development process. The system’s detectable disorder coverage is comparable to that of a human expert. Additionally, the system’s excellent generalizability and explainability enhance its clinical applicability.

### Annotated and disorder-contained data

Most existing deep-learning medical systems rely on supervised learning, which requires a substantial amount of annotated data to achieve generalizability, accuracy, and recognition gratuity. However, obtaining sufficient annotated data in medical image research is challenging due to the time-consuming and expert knowledge-intensive nature of the notating process. For instance, even for an experienced expert, it may take several minutes to annotate a medical image at the region-level, which provides strong supervision for disorder detection by indicating the exact lesion region. Consequently, research works that rely on region-level annotation, such as Nikolov et al.^7^ and Monteiro et al.,^8^ are limited by the amount of annotated data, which hinders the generalizability and accuracy of the system.

To reduce the dependence on annotated data, researchers have explored alternative learning strategies for medical image research. For instance, weakly-supervised learning^9,10,11^ allows each training sample to lack a label or have an incorrect label, significantly reducing the annotation cost for experts. Unsupervised learning, on the other hand, uses unannotated training data to enhance the feature representation capacity of a deep learning network, thereby reducing the number of required annotated samples. Self-supervised learning is a recent representative unsupervised learning method^12, 13^ that annotates each sample by itself instead of relying on human experts. However, these learning strategies require a substantial amount of disorder-contained data to ensure accuracy. Collecting enough disorder-contained data is challenging for general researchers due to ethical and legal considerations, limiting related research to large medical institutions. For example, Chilamkurthy et al.^5^ collected over 300,000 brain CT scans from more than 20 medical centers, which is beyond the reach of most researchers.

Compared to previously adopted learning strategies in medical image research, the proposed ISL tackles a challenging task where no annotated or disorder-contained data is available. The only available data is disorder-free data, which can be easily obtained by any medical institution capable of medical imaging scans.

### Disorder coverage

The clinical application of medical image research is an important goal. However, most existing works focus on only one or two common disorder types,^5, 14^ even for systems developed by institutions with abundant medical resources. For instance, the system^5^ is derived from over 300,000 scans, yet it can only recognize four types of disorders. This challenge arises from two aspects. Firstly, it is impractical for researchers to construct models for each disorder type due to the difficulty of collecting and annotating medical images. Secondly, developing models for rare disorders with previous learning strategies is challenging when only a few samples are available. With ISL, researchers do not need to collect data or construct models for specific disorders, enabling the built system to achieve broad-spectrum disorder detection.

### Anomaly Detection

Distinguishing disorder-contained images from disorder-free ones can be viewed as an anomaly detection problem, which is a popular research field in machine learning. An intuitive assumption is that anomalies lie outside the distribution of normal samples. Therefore, it is natural to train a classifier to differentiate abnormal samples from normal ones.^15,16^

Recent works have utilized generation networks for anomaly detection, employing two primary approaches: (1) utilizing the latent feature;^17, 18, 19^ and (2) utilizing the reconstructed image.^20, 21, 22^ In the first approach, a generation network produces a latent feature and a reconstructed image when an image is inputted. The latent features can be used to determine whether the image is abnormal. In the second approach, a generation network produces a corresponding normal image for a given image. If the original image contains abnormal characteristics, it can be recognized based on the difference between the original and generated images. In the field of medical image analysis, two types of methods have achieved certain results in specific diseases.^23,24,25^ For instance, Yao et al.^23^ used the second approach to generate healthy pulmonary CT images, which were used to determine whether the lungs contained COVID-19.

However, both approaches have limitations when applied to broad-spectrum disorder detection in medical images. In the first approach, medical images are complex, which results in complex latent features. Therefore, recognizing disorder-contained images based solely on latent features is challenging. To demonstrate this, we compared our method with a baseline method that directly fed original medical images into the disorder recognition network. The baseline method achieved an average AUC of 0.653 on the prospective dataset, which is inferior to the results (AUC 0.868) obtained by our method. In the second approach, existing generation-based methods reconstruct the original disorder tissues of a medical image due to the strong feature representation capability of generative networks, which fails to achieve abnormal recognition. With the generation strategy in ISL, only context images and global structure information are provided, allowing the generation network to eliminate the interference of the original disorder tissue and conceive healthy tissues like a radiologist. To showcase the efficacy of our system in comparison to existing techniques, we conducted a comparative analysis with other representative reconstruction-based anomaly detection methods, specifically Auto-Encoder,^26^ AnoGAN,^17^ GANomaly,^27^ pix2pix,^28^ and Cycle-GAN.^29^ The experiment was carried out on the task of detecting pulmonary disorders. The results of the analysis are presented in Supplementary Table 12. Our system outperformed the baselines, with the highest AUC of 0.846 achieved by GANomaly, which is 0.047 lower than our method. This significant improvement underscores the ability of our ISL-based system to successfully accomplish disorder recognition tasks.

## Methods

### CT scan collection

Initially, we retrieved 954,508 scans from the PACS of the PLAGH between March 2012 and July 2019. These scans contained CT images stored in DICOM (digital imaging and communications in medicine) format, and all DICOMs were de-identified before data analysis. We then screened the scans by excluding reconstructed scans (processed with algorithms in CT machines), non-axial-section scans (coronal section and sagittal section scans), non-head scans (scans of breast and full-body, etc.), and non-origin scans (scans of CTA and CTP, etc.). The inclusion and exclusion criteria of the screening process are detailed in Figure 8. After screening, a total of 62,239 CT scans with an average slice number of 28 were selected.

**Figure 8.**
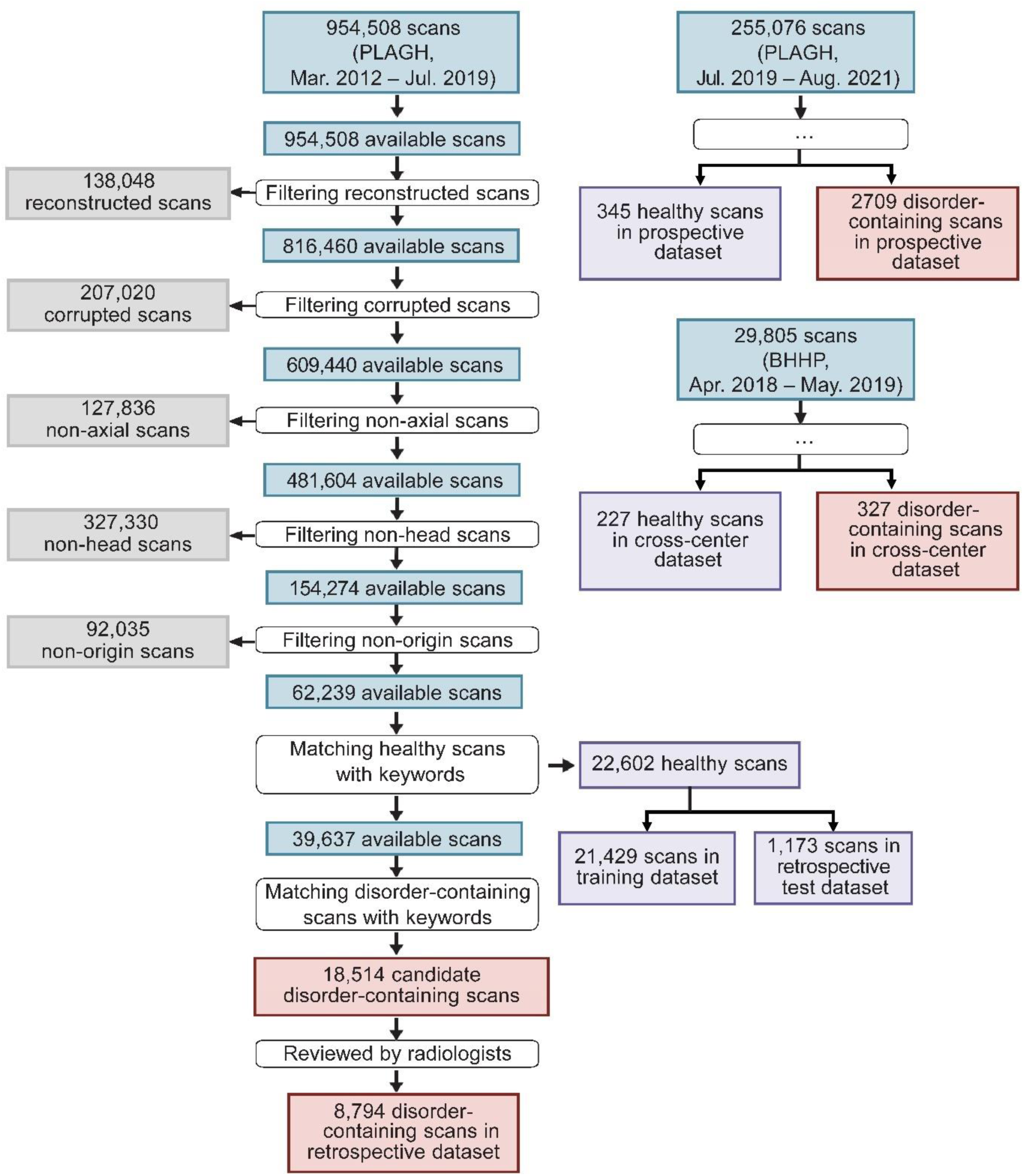
The process of constructing the datasets. *Left*, 954,508 scans (March 2012 - July 2019) were collected from the PLAGH by retrieving head CT-related keywords. After a series of filtering steps, a training dataset and a retrospective dataset were constructed. The trainingdataset consisted of 21,429 healthy scans, and the retrospective dataset consisted of 1,173 healthy scans and 8,794 disorder-containing scans. *Right*, we collected 255,076 scans (July 2019 - August 2021) from the PLAGH and 29,805 scans (April 2018 - May 2019) from the BHHP. We used these data to construct the prospective and cross-center test datasets using the same process.

### Disorder types statistics

Each retrieved scan includes a clinical report written by an interpreting radiologist during the examination. To determine the disorder types to be interpreted by our system, we first applied a rule-based NLP algorithm to the clinical reports. This algorithm counted the occurrence frequencies of different word phrases. We then invited three radiologists to analyze the frequency statistics results and select the disorder types to be evaluated. Ultimately, 127 types of disorders were selected.

Our method selection was primarily driven by the desire to create a system capable of handling the most common types of disorders encountered in actual clinical practice, while also ensuring a comprehensive coverage of various disorder types. Our dataset, which spans nearly seven years (2012 to 2019) and includes data from 301 hospitals, is believed to encompass most types of disorders. By focusing on the most frequently occurring disorder types, we aimed to enhance the practicality of our system, ensuring it is well-equipped to manage common disorders while maintaining a broad scope of disorder types.

### Training dataset and test dataset

The construction of the development and test datasets relied on the clinical reports, which were considered the gold standard. The training dataset only included disorder-free CT scans, and their reports uniformly described them as “No abnormality is observed.” Therefore, we could efficiently obtain disorder-free CT scans. Ultimately, we selected 22,602 disorder-free scans, which were divided into two parts: the training dataset (21,429 scans) and the negative samples in the retrospective test dataset (1,173 scans), detailed data statistics are shown in Supplementary Table 5.

Regarding the positive samples (disorder-contained scans) in the test dataset, we initially retrieved 18,514 scans using stated disorder-related keywords. We then invited 30 board-certified radiologists with 6 to 15 years of experience to label each scan based on the images and its associated report. The radiologists assigned a binary label (i.e., 0, 1) to each scan, where 1 indicated that the scan contained the expected disorder type. Ultimately, 8,794 scans were labeled as 1 and selected as the positive samples in the retrospective test dataset. The prospective and cross-center test datasets were constructed similarly. However, due to the smaller data amounts compared to the retrospective dataset, they also contained a smaller number of disorder types, specifically 116 and 46, respectively. Please refer to Supplementary Table 9-11 for the detailed data statistics.

### Developing a system with inversed-supervised learning

ISL allows for the training of a deep learning network without accessing disorder-contained samples, enabling researchers with only general and healthy images to build a broad-spectrum disorder detection system. ISL is built on two technologies: missing information completion and data distribution estimation. Missing information completion enables a system to reconstruct healthy tissues of masked parts of a medical image using a de-disorder network (DeDN) derived from general and healthy images. The scanning medical images of the human body are relatively standardized. Therefore, for healthy images, the reconstructed version should be very close to the original version. And for a medical image containing any disorders, the reconstructed image will differ significantly from the original. Data distribution estimation requires the estimation of the distribution of healthy difference images, which are calculated using healthy images and their reconstructed images generated by a DeDN. If an image contains any disorders, its difference image will fall outside the distribution and be detected. Notably, unlike many existing disorder detection algorithms, ISL can detect a significantly increased number of disorder types.

### CT slice conversion

In our dataset, the pixel values in a CT scan are represented by 14-bit numbers, which exceed the range of human perception. To address this, we converted each CT slice into a 3-channel 8-bit image, conforming to the standard image format and suitable for display. Radiologists typically use specific window locations (WL) and window widths (WW) to observe various types of disorders. Building on this, we applied specific WL and WW settings for the image conversion, with the specific settings outlined in Supplementary Table 16.

### Problem Formulation

ISL is designed to address binary-classification problems by predicting the probability of the presence of any disorder type in a medical image. For example, in brain CT scans, the input of an ISL-based system is a slice **x***_i_* from a brain CT scan for a CT scan 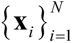, where *N* is the slice number of this scan. The system output *p_i_* indicates the probability of any disorder type in the slice **x***_i_*. During deployment, the slice-level outputs 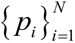 are aggregated to a scan-level output *p*’ by averaging the probabilities of all the slices in the scan, where 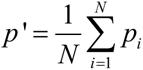. We adopt the slice-wise processing method because we believe that, for the initial assessment of disorders in medical image analysis, the information provided by a single image is already adequate. Slice-wise processing offers a more efficient strategy, where sequential information is utilized to confirm the precise categories of disorders. As the ISL-based system processes individual slices, we have omitted the subscript number of slices in a scan for the sake of conciseness in the following method introduction.

An ISL-based system comprises two networks: a de-disorder network (DeDN) and a disorder recognition network (DRN). Given a medical image **x**, we first use a DeDN to generate a de-disorder image **x***_dd_* of **x**. If **x** contains a disorder, the disorder tissues in the area are converted into healthy ones. Then, the difference image **x***_dif_* = ∥**x** − **x***_dd_*∥ is input into the DRN network, which predicts the probability *p* of disorder containment in the image. The overview of ISL is shown in Figure 9.

**Figure 9.**
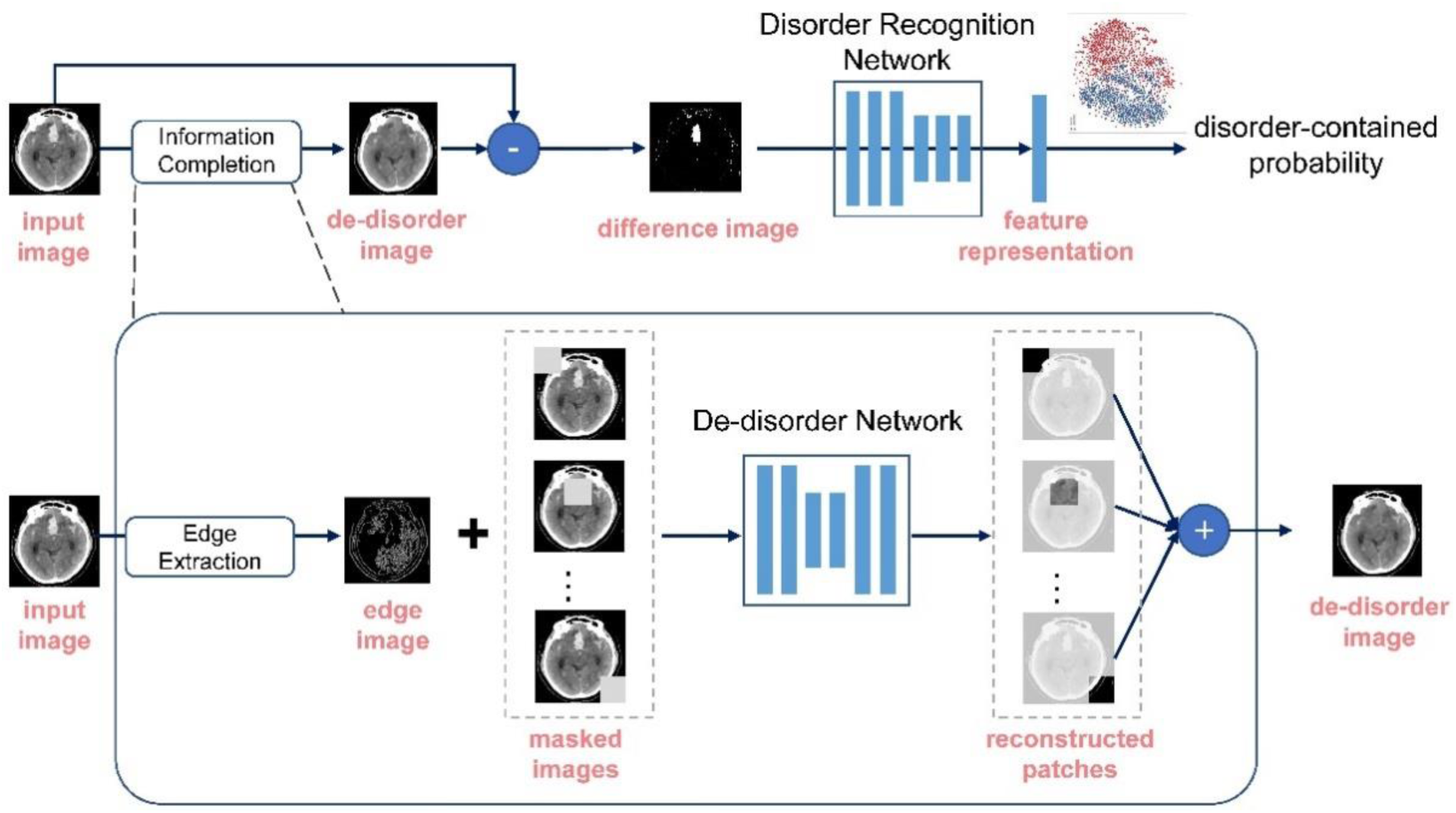
The overview of ISL, a learning algorithm for developing broad-spectrum disorder detection systems. The training dataset consists only of healthy scans, and a de-disorder network is learned to generate de-disorder images. A disorder recognition network is then employed to predict the probability of disorder containment based on the difference image obtained by subtracting the input and de-disorder image. This approach enables the developed model to achieve broad-spectrum disorder detection even without any disorder-contained data.

### De-disorder network

Given a masked image, **x̅**=**x**·**m**, where **m** is an image mask of **x**, a deep encoder-decoder network (DeDN) can predict the masked region and generate a reconstructed image **x̂**. The detailed architecture of the DeDN is shown in Supplementary Table 15, which has been proven to be effective in many image generation tasks. In our architecture comparison experiment (Supplementary Table 13), we found that the adopted architecture has already captured the most salient features necessary for generating high-quality medical images. In this study, we utilized the DeDN to generate de-disordered medical images. Specifically, we divided a medical image **x** into *K* × *K* grids of uniform size. For each grid with coordinates (*i*, *j*), where 1 ≤ *i* ≤ *K* and 1 ≤ *j* ≤ *K*, we applied a mask **m**^(*i,j*)^ to erase it and obtain a masked image. The DeDN was then used to reconstruct the masked image and generate the reconstructed image **x̂**^(*i, j*)^ Finally, we combined the *K*×*K* generated images into a reconstructed de-disordered medical image using the following equation:

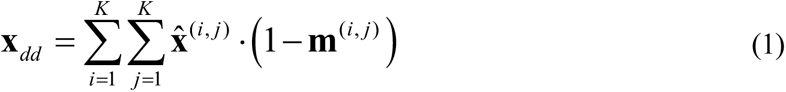

We will use **x***_dd_* for comparative analysis with the original image **x**. A deep encoder-decoder network (DeDN) takes as input a masked image **x̅**^(*i, j*)^ and multiple image edge maps 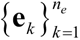 of **x**. Edge maps retain structural information of the masked region, which can improve the quality of the reconstructed image. Edge maps can be constructed using mature image processing schemes, such as the Canny Edge Detector.^30^ The DeDN *G* generates the reconstructed image, **x̂**^(*i, j*)^ using the following equation:

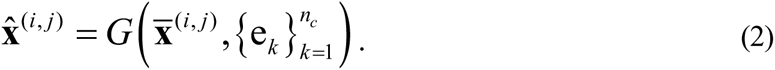

We trained the network using a joint loss:

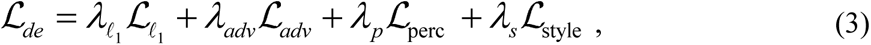

which includes an *ℓ*_1_ loss, adversarial loss, perceptual loss, and style loss. The *ℓ*_1_ loss minimizes the reconstruction error between **x̂**^(*i, j*)^ and **x**. The adversarial loss *ℒ_adv_* ensures the reality of the generated image and is defined as:

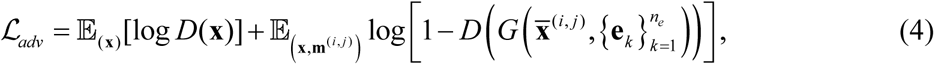

where *D* is the discriminator network. We also included perceptual loss *ℒ_prec_* and style loss *ℒ_style_*, following Nazeri et al.^31^ The perceptual loss *ℒ_prec_* penalizes reconstructed images that are not perceptually similar to the original ones and is defined as a distance measure between activation maps of a pretrained network:

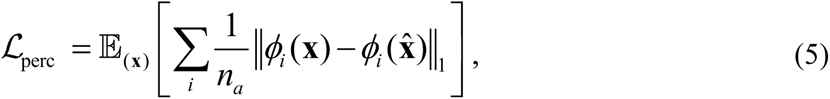

where *ϕ_i_* is the activation map of the *i*^th^ layer of the pretrained VGG-19 network, and *n_a_* is the number of layers. We chose the output of the first ReLU activation layer in each of the five blocks^31^ of VGG-19 pretrained on the ImageNet dataset.^32^ This choice was based on its proven effectiveness in capturing image features. The comparison experiment on the cross-center test dataset showed similar results to ResNet34 (Supplementary Table 14), indicating the robustness of our model to the choice of architecture for perceptual loss calculation.

The style loss is calculated based on these activation maps and is an effective tool to alleviate the “checkerboard” artifacts caused by transpose convolution layers. The loss measures the differences between covariances of the activation maps and is defined as:

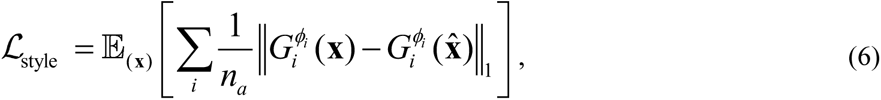

where 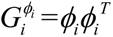 is a Gram matrix constructed from the activation map *ϕ_i_*. The Gram matrix of an activation map captures the correlation between different channels and the texture structure of its corresponding image. For a real medical image, the Gram matrix resembles the identity matrix, with larger diagonal values indicating strong correlations within the same feature and smaller off-diagonal values reflecting feature in-dependence. Conversely, a blurry and texture-lacking generated image results in a constant Gram matrix with similar values for each element, indicating a lack of feature differentiation. To optimize the model, we minimize the difference between the Gram matrices of the real and generated images.

### Disorder recognition network

To minimize the impact of reconstructed noise on disorder detection and improve the performance further, we developed a disorder recognition network that takes the difference image **x***_dif_* as input and extracts an embedded representation in the latent space. We designated the embedding distribution of difference images from disorder-free data as the reference distribution. The disorder recognition network should ensure that the embeddings of disorder-free data are centralized and compact, while the embeddings of disorder-contained data are random and as far as possiblefrom the reference distribution. In this case, the distance between an embedded representation and the center of the reference distribution can effectively indicate the possibility of disorder presence.

Inspired by the support vector data description (SVDD) algorithm^33^ and the contrastive learning method,^34^ we developed the DRN based on augmentation views. In addition to a given healthy medical image **x**, the DRN uses two augmented views, **x**^−^ and **x**^+^, generated from **x** for network training. **x**^−^ is produced with rotation and flipping transformations, yielding an appearance akin to **x**. **x**^+^, on the other hand, is generated with cutout transformation, which can damage healthy tissues in **x**. As a result, **x**^+^ disrupts the inherent distribution of the healthy image **x** and is thus considered a disorder-contained view. For DRN, the main basis for judgment is the size of the pixel difference and the range of difference. Therefore, by applying cutout transformation to the original healthy image, we can obtain images with large pixel differences and a wide range of differences. This difference, or ‘anomaly’, is what DRN is trained to detect.

After training the DeDN, the input of the DRN consists of three parts: the reference difference image **x***_dif_*, the negative difference image 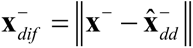, and the positive difference image 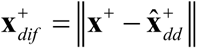, where **x̂**^+^ and **x̂**^−^ denote the reconstructed images of **x**^−^ and **x**^+^, respectively. The DRN extracts embedded representations of the difference images, denoted as **h**, = **h**^−^, and **h**^+^. The DRN learns reasonable embedded representations of disorder-free input by maximizing the similarity between **h**^−^ and **h** while distinguishing **h**^+^ from **h**. In this study, we used the Euclidean distance as the metric to measure the similarity of the embeddings. We pretrained the network using MoCo^12^ and averaged the embeddings of the wide-ranging training dataset. The averaged embedding **c** is considered the center of the reference distribution. Setting the center as an anchor, we designed a compactness loss to maximize the similarity between the negative embeddings:

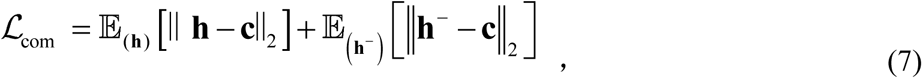

Where *ℒ_com_* minimizes the distances between the embeddings **h**, **h**^−^ and the reference center, which ensures that the DRN can extract consistent features for disorder-free difference images. To further improve the discriminative ability of the network, we used a discrimination loss *ℒ_dis_*, whichforces the network to maximize the distance between the reference center and the embedded representation of **x**^+^ :

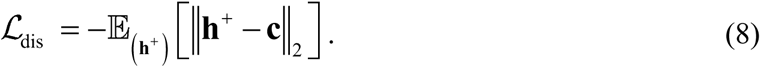

The overall loss function utilized to train the DRN is defined as:

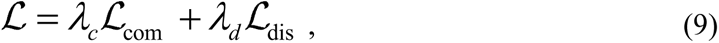

where *λ_c_* and *λ_d_* are the weights of the loss functions *ℒ_com_* and *ℒ_dis_*, respectively.

### Disorder visualization

The ISL-based system is capable of identifying the locations of disorders, which is crucial for clinical applications.^35, 36, 1^ Higher pixel values in the image regions of **^x^***_dif_* indicate a higher likelihood of the presence of a disorder. To enhance the visual appeal of the results, we conducted several post-processing steps on **x***_dif_* : (1) Eliminating the bias caused by the normal range reconstruction. Pixels with values below a certain threshold *t* were set to zero. (2) Reducing reconstruction noise. After normalizing the pixel values to the range of [0, 1], weadded the values of the *s* × *s* region surrounding each pixel to itself. This smoothing technique reduced the noise in the image. (3) Enhancing the disorder area. We utilized an exponential function to manipulate the pixel values, resulting in an amplification of the differences in values among pixels. This process serves to accentuate the presence of disorder within the region of interest.

With processed **x*_dis_***, which is denoted as 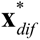, we employed the Average Pixel Difference Score (APDS) as a metric to quantify the discrepancy between the original and the reconstructed images. The APDS is computed by averaging the pixel values of the processed **x***_dif_*, within the effective pixel area. This area encompasses human body structures and is differentiated by non-zero pixel values. Formally, given an image **x**, its APDS is calculated as the ratio of the sum of pixel values in the original image **x** to the number of non-zero pixels in the corresponding processed difference image 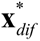, denoted as 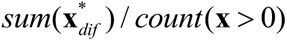. Our experimental results revealed that the APDS for normal images was approximately 5 × 10^−5^. In contrast, for images with lesions, this metric typically escalated to an order of 5 × 10^−4^. The observed difference in these metrics is substantial enough to effectively distinguish between normal images and those with lesions.

### Model selection and statistical analysis

Since we were unable to access data containing disorders for model evaluation during training, we selected the model when the training loss did not decrease for 5 consecutive epochs. The primary parameters of our system, including the network architectures, hyperparameter values, and optimization strategies, are presented in Supplementary Tables 15 and 16. To ensure statistical significance, we applied 95% confidence interval (CI). Specifically, for each iteration, we randomly sampled 30% of CT scans from the test dataset for evaluation. We repeated this procedure 1,000 times and calculated the 95% CI of the evaluation metrics for the model. To determine the optimal classification threshold, we used a derivative-based method, specifically by maximizing the harmonic mean of sensitivity and specificity. This is expressed in the following optimization criterion: [Maximize (2 * sensitivity * specificity) / (sensitivity + specificity)]. This criterion is known as a variant of the F1 score, which balances sensitivity and specificity to achieve the best trade-off between the two.

### Code and software availability

The system was developed using standard libraries and scripts available in Porch. The developing code is at https://gitlab.com/heyuwei403/islcode. The code will be made publicly available after the acceptance. A demo video of our diagnosis and visualization software is at https://gitlab.com/heyuwei403/isl-system-demo.

### Data availability

The datasets to develop our head and pulmonary disorder detection system are not publicly available due to the privacy requirement of the PLAGH and FAHGMU. The cross-center dataset (554 scans and experts’ annotations) from BHHP is allowed to be distributed for research purposes from the corresponding author upon reasonable request. The development and evaluation dataset for retinal OCT disorder detection can be downloaded from https://data.mendeley.com/ *datasets/rscbjbr9sj/2*.

## Acknowledgements

We thank the radiologists from the PLAGH for their efforts in labeling the test data. We thank National Key R&D Program of China (2020AAA0105500 to Y.G., G.D., and Q.D., 2018YFA0704000 to F.X.), and National Natural Science Foundation of China (U21B2013 to Y.G., 62021002 to F.X., 81825012 to X.L., 81730048 to X.L., 82271952 to J.L.) for supporting this study.

## Author’s contributions

X.L., Q.D., F.X., Y.H. and Y.G. designed the research; X.L., J.L., Y.H., W.Z. and H.L. collected the data; Y.H., Y.G., L.M., J.L., X.L. and F.X. verified the raw underlying data, which had been accessed by all authors; Y.H., Y.G., L.M. and H.T. developed the system; Y.H., Y.G., L.M., G.D., J.L., H.L. and J.H. analyzed the results; Y.H., L.M. and J.L. co-wrote the manuscript; S.L., H.Q., F.X. and Y.G. critically revised the manuscript; and all the authors discussed the results and provided feedback regarding the manuscript.

