## Supplementary material for "Disorder-Free Data are All You Need: Inverse Supervised Learning for Broad-Spectrum Head Disorder Detection": Supplemenal tables and figures

### Table of Contents

1 **Supplementary Table 1. Performance on the retrospective dataset for head disorder detection.** 3

2 **Supplementary Table 2. Performance on the prospective dataset for head disorder detection.** 6

3 **Supplementary Table 3. Performance on the cross-center dataset for head disorder detection.** 9

4 **Supplementary Table 4. True positive rate (TPR) at different false positive rate (FPR) levels.** 10

5 **Supplementary Table 5. Data characteristics of healthy scans in the training dataset, as well as those in the prospective, retrospective, and cross-center test datasets**…………………………...11

9 **Supplementary Table 9. Statistics of the disorder-containing scans in the retrospective dataset………** ........................................................................................................................ 14

10 **Supplementary Table 10. Statistics of the disorder-containing scans in the prospective dataset………………** ............................................................................................................ 17

11 **Supplementary Table 11. Statistics of the disorder-containing scans in the cross-center dataset.** ……………………………………………………………………………………………...20

**Supplementary Table 1. Performance on the retrospective dataset for head disorder detection.**

| | Retrospective | | | |
|---|---|---|---|---|
| | **Num** | **AUC** | **Sensitivity** | **Specificity** |
| Effusion | 487 | 0.963 (0.952, 0.975) | 0.920 (0.897, 0.945) | 0.890 (0.872, 0.907) |
| Subarachnoid Hemorrhage | 485 | 0.976 (0.967, 0.985) | 0.928 (0.905, 0.953) | 0.922 (0.906, 0.938) |
| Subdural Hematoma | 482 | 0.958 (0.946, 0.971) | 0.898 (0.873, 0.925) | 0.890 (0.872, 0.909) |
| Pneumocephaly | 474 | 0.967 (0.956, 0.979) | 0.922 (0.899, 0.947) | 0.915 (0.899, 0.933) |
| Parenchymal Hemorrhage | 474 | 0.955 (0.943, 0.969) | 0.901 (0.876, 0.928) | 0.890 (0.873, 0.908) |
| Multiple Cerebral Infarctions | 465 | 0.865 (0.844, 0.887) | 0.738 (0.699, 0.776) | 0.866 (0.847, 0.886) |
| Corona Radiata Cerebral Infarction | 459 | 0.667 (0.636, 0.698) | 0.560 (0.514, 0.606) | 0.688 (0.662, 0.716) |
| Lacunar Infarction | 456 | 0.687 (0.657, 0.719) | 0.667 (0.623, 0.713) | 0.596 (0.568, 0.624) |
| Basal Ganglia Ischemia | 454 | 0.861 (0.839, 0.885) | 0.731 (0.692, 0.773) | 0.865 (0.847, 0.885) |
| Basal Ganglia Cerebral Infarction | 453 | 0.716 (0.687, 0.747) | 0.561 (0.512, 0.609) | 0.778 (0.754, 0.802) |
| Ischemia | 444 | 0.928 (0.914, 0.945) | 0.867 (0.838, 0.899) | 0.837 (0.816, 0.858) |
| Calcification | 442 | 0.825 (0.801, 0.852) | 0.692 (0.647, 0.738) | 0.836 (0.814, 0.858) |
| Malacia Foci | 436 | 0.917 (0.900, 0.935) | 0.853 (0.821, 0.890) | 0.833 (0.811, 0.856) |
| Contusion | 424 | 0.954 (0.940, 0.969) | 0.906 (0.880, 0.934) | 0.897 (0.879, 0.914) |
| Cerebral Infarction Of Caudate Nucleus Head | 422 | 0.898 (0.878, 0.916) | 0.820 (0.784, 0.855) | 0.813 (0.790, 0.836) |
| Periventricular Cerebral Infarction | 397 | 0.803 (0.777, 0.829) | 0.713 (0.668, 0.758) | 0.733 (0.707, 0.759) |
| Tubercle | 362 | 0.819 (0.792, 0.846) | 0.671 (0.624, 0.721) | 0.830 (0.810, 0.852) |
| Intraventricular Hemorrhage | 323 | 0.986 (0.978, 0.995) | 0.944 (0.923, 0.969) | 0.942 (0.929, 0.957) |
| Brainswelling | 315 | 0.952 (0.938, 0.969) | 0.898 (0.867, 0.933) | 0.880 (0.862, 0.900) |
| Sclerosis | 272 | 0.840 (0.810, 0.872) | 0.746 (0.695, 0.801) | 0.823 (0.799, 0.844) |
| Space Occupying Lesions | 261 | 0.925 (0.904, 0.946) | 0.862 (0.820, 0.904) | 0.847 (0.827, 0.869) |
| Epidural Hematoma | 252 | 0.947 (0.927, 0.967) | 0.893 (0.857, 0.929) | 0.892 (0.875, 0.912) |
| Cerebral Edema | 216 | 0.965 (0.952, 0.981) | 0.903 (0.866, 0.944) | 0.900 (0.882, 0.918) |
| Minor Hemorrhage | 212 | 0.966 (0.953, 0.982) | 0.910 (0.873, 0.953) | 0.892 (0.874, 0.911) |
| Thalamic Cerebral Infarction | 205 | 0.713 (0.671, 0.754) | 0.610 (0.546, 0.673) | 0.690 (0.662, 0.717) |
| Soft Tissue Swelling | 203 | 0.937 (0.916, 0.962) | 0.877 (0.833, 0.926) | 0.852 (0.831, 0.873) |
| Arteriosclerosis | 196 | 0.810 (0.771, 0.849) | 0.668 (0.602, 0.735) | 0.831 (0.809, 0.853) |
| Parenchymal Hematoma | 176 | 0.982 (0.971, 0.995) | 0.949 (0.920, 0.983) | 0.938 (0.924, 0.952) |
| Centrum Semiovale Cerebral Infarction | 151 | 0.733 (0.690, 0.781) | 0.702 (0.636, 0.775) | 0.663 (0.635, 0.693) |
| Parietal Lobe Cerebral Infarction | 148 | 0.788 (0.742, 0.836) | 0.622 (0.541, 0.703) | 0.874 (0.856, 0.894) |
| Frontal Lobe Cerebral Infarction | 123 | 0.729 (0.674, 0.787) | 0.537 (0.447, 0.626) | 0.880 (0.862, 0.899) |
| Arachnoid Cyst | 119 | 0.844 (0.806, 0.886) | 0.714 (0.639, 0.798) | 0.831 (0.809, 0.852) |
| Hydrocephalus | 108 | 0.999 (0.998, 1.000) | 0.991 (0.981, 1.000) | 0.969 (0.961, 0.980) |
| Cerebral White Matter Degeneration | 107 | 0.831 (0.787, 0.878) | 0.682 (0.598, 0.776) | 0.851 (0.830, 0.871) |
| Paraventricular Ischemia | 104 | 0.875 (0.834, 0.921) | 0.702 (0.615, 0.798) | 0.938 (0.925, 0.953) |
| Cavum Septum Pellucidum | 102 | 0.842 (0.801, 0.887) | 0.814 (0.745, 0.892) | 0.689 (0.662, 0.716) |
| Subcutaneous Hematoma | 102 | 0.896 (0.857, 0.936) | 0.843 (0.775, 0.912) | 0.816 (0.793, 0.837) |
| Temporal Bone Fracture | 101 | 0.915 (0.875, 0.959) | 0.871 (0.812, 0.941) | 0.831 (0.809, 0.854) |
| Frontal Lobe Ischemia | 92 | 0.842 (0.795, 0.893) | 0.815 (0.739, 0.891) | 0.753 (0.729, 0.778) |
| Subdural Hemorrhage | 89 | 0.985 (0.971, 1.000) | 0.955 (0.921, 1.000) | 0.943 (0.930, 0.957) |
| Ventriculomegaly | 83 | 0.992 (0.985, 1.000) | 0.976 (0.952, 1.000) | 0.817 (0.795, 0.839) |
| Parietal Fracture | 83 | 0.909 (0.864, 0.960) | 0.880 (0.807, 0.952) | 0.831 (0.809, 0.854) |
| Occipital Fracture | 82 | 0.922 (0.881, 0.971) | 0.878 (0.817, 0.951) | 0.867 (0.847, 0.886) |
| Occipital Lobe Cerebral Infarction | 73 | 0.918 (0.877, 0.962) | 0.849 (0.767, 0.932) | 0.841 (0.819, 0.862) |
| Frontal Bone Fracture | 71 | 0.875 (0.821, 0.936) | 0.817 (0.732, 0.915) | 0.760 (0.736, 0.784) |

| Retrospective | | | | |
|---|---|---|---|---|
| | Num | AUC | Sensitivity | Specificity |
| Osteoma | 70 | 0.844 (0.790, 0.906) | 0.743 (0.643, 0.843) | 0.865 (0.845, 0.886) |
| Temporal Lobe Cerebral Infarction | 66 | 0.915 (0.880, 0.958) | 0.848 (0.773, 0.939) | 0.829 (0.807, 0.851) |
| Old Cerebral Infarction | 66 | 0.822 (0.759, 0.890) | 0.636 (0.515, 0.758) | 0.906 (0.889, 0.923) |
| Lipoma | 63 | 0.887 (0.834, 0.943) | 0.841 (0.762, 0.937) | 0.753 (0.728, 0.779) |
| Scalp Hematoma | 63 | 0.907 (0.855, 0.968) | 0.873 (0.794, 0.968) | 0.837 (0.816, 0.858) |
| Artifacts Shadow | 62 | 0.969 (0.944, 1.000) | 0.935 (0.887, 1.000) | 0.866 (0.846, 0.886) |
| Brain Atrophy | 59 | 0.953 (0.925, 0.989) | 0.915 (0.847, 1.000) | 0.805 (0.781, 0.829) |
| Cisterna Magna | 59 | 0.870 (0.824, 0.921) | 0.847 (0.763, 0.949) | 0.719 (0.693, 0.746) |
| Herniation | 55 | 0.999 (0.999, 1.000) | 1.000 (1.000, 1.000) | 0.969 (0.960, 0.980) |
| Skull Fracture | 54 | 0.937 (0.894, 0.990) | 0.889 (0.815, 0.981) | 0.865 (0.846, 0.885) |
| Absence Of Bone | 50 | 0.976 (0.960, 0.996) | 0.940 (0.880, 1.000) | 0.890 (0.872, 0.909) |
| Hemispheric Infarction | 50 | 0.980 (0.962, 1.000) | 0.940 (0.880, 1.000) | 0.922 (0.907, 0.938) |
| Brainstem Infarction | 48 | 0.890 (0.844, 0.939) | 0.875 (0.792, 0.979) | 0.760 (0.735, 0.786) |
| Multiple Hemorrhages | 47 | 0.945 (0.903, 0.997) | 0.915 (0.851, 1.000) | 0.892 (0.875, 0.911) |
| Metal Shadow | 40 | 0.948 (0.912, 0.991) | 0.875 (0.775, 0.975) | 0.844 (0.823, 0.865) |
| Metastases | 40 | 0.936 (0.902, 0.974) | 0.850 (0.750, 0.975) | 0.801 (0.777, 0.825) |
| Centrum Semiovale Ischemia | 39 | 0.845 (0.771, 0.928) | 0.718 (0.590, 0.872) | 0.846 (0.825, 0.867) |
| Atherosclerosis | 38 | 0.948 (0.909, 0.995) | 0.895 (0.816, 1.000) | 0.819 (0.797, 0.842) |
| Midline Shift | 37 | 0.941 (0.894, 1.000) | 0.892 (0.811, 1.000) | 0.880 (0.862, 0.900) |
| Facial Bone Fracture | 31 | 0.990 (0.981, 1.000) | 0.968 (0.935, 1.000) | 0.893 (0.876, 0.912) |
| Corona Radiata Ischemia | 31 | 0.895 (0.844, 0.956) | 0.871 (0.774, 1.000) | 0.774 (0.749, 0.800) |
| Leukoaraiosis | 26 | 0.893 (0.831, 0.973) | 0.846 (0.731, 1.000) | 0.788 (0.764, 0.811) |
| Insular Infarction | 26 | 0.965 (0.943, 0.993) | 0.923 (0.846, 1.000) | 0.876 (0.858, 0.895) |
| Parietal Lobe Ischemia | 22 | 0.886 (0.811, 0.985) | 0.909 (0.818, 1.000) | 0.744 (0.719, 0.769) |
| Cerebellar Hemisphere Infarction | 21 | 0.894 (0.830, 0.968) | 0.857 (0.714, 1.000) | 0.771 (0.747, 0.795) |
| Minor Haematoma | 21 | 0.918 (0.849, 1.000) | 0.905 (0.810, 1.000) | 0.884 (0.865, 0.902) |
| Osseous Defects | 20 | 0.947 (0.904, 1.000) | 0.950 (0.900, 1.000) | 0.893 (0.876, 0.913) |
| Lacunar Ischemia | 19 | 0.935 (0.878, 1.000) | 0.895 (0.789, 1.000) | 0.894 (0.876, 0.912) |
| Zygomatic Fracture | 17 | 0.889 (0.797, 1.000) | 0.824 (0.647, 1.000) | 0.757 (0.733, 0.781) |
| Widened Cisterna Magna | 16 | 0.940 (0.907, 0.975) | 1.000 (1.000, 1.000) | 0.831 (0.809, 0.853) |
| Meningioma | 16 | 0.943 (0.900, 1.000) | 0.938 (0.875, 1.000) | 0.754 (0.730, 0.781) |
| Tumour | 16 | 0.970 (0.947, 0.999) | 1.000 (1.000, 1.000) | 0.825 (0.803, 0.846) |
| Widened Subarachnoid Space | 15 | 0.907 (0.825, 1.000) | 0.933 (0.867, 1.000) | 0.841 (0.819, 0.862) |
| Sphenoid Fracture | 14 | 0.931 (0.869, 0.999) | 0.929 (0.857, 1.000) | 0.725 (0.698, 0.750) |
| Malformation | 13 | 0.992 (0.985, 1.000) | 1.000 (1.000, 1.000) | 0.902 (0.885, 0.920) |
| Cystic Foci | 13 | 0.835 (0.699, 1.000) | 0.846 (0.692, 1.000) | 0.637 (0.609, 0.667) |
| Sella Turcica Enlargement | 12 | 0.923 (0.875, 0.978) | 1.000 (1.000, 1.000) | 0.773 (0.748, 0.796) |
| Choroid Cyst | 12 | 0.759 (0.616, 0.917) | 0.833 (0.667, 1.000) | 0.586 (0.556, 0.615) |
| Septum Pellucidum Cyst | 12 | 0.743 (0.597, 0.917) | 0.750 (0.500, 1.000) | 0.620 (0.592, 0.649) |
| Wallerian Degeneration | 11 | 0.973 (0.946, 1.000) | 1.000 (1.000, 1.000) | 0.760 (0.737, 0.786) |
| Senile Encephalopathy | 11 | 0.926 (0.851, 1.000) | 1.000 (1.000, 1.000) | 0.265 (0.239, 0.289) |
| Enhancement Foci | 11 | 0.950 (0.901, 1.000) | 0.909 (0.818, 1.000) | 0.878 (0.858, 0.898) |
| Skull Base Fracture | 11 | 0.981 (0.962, 1.000) | 1.000 (1.000, 1.000) | 0.837 (0.815, 0.858) |

| Retrospective | | | | |
|---|---|---|---|---|
| | **Num** | **AUC** | **Sensitivity** | **Specificity** |
| Zygomatic Arch Fracture | 11 | 0.870 (0.741, 1.000) | 0.818 (0.636, 1.000) | 0.757 (0.732, 0.781) |
| Epidural Hemorrhage | 10 | 0.936 (0.873, 1.000) | 1.000 (1.000, 1.000) | 0.455 (0.427, 0.483) |
| Fat Deposition | 9 | 0.969 (0.943, 1.000) | 1.000 (1.000, 1.000) | 0.878 (0.860, 0.898) |
| Fat Density | 9 | 0.858 (0.745, 0.992) | 0.778 (0.556, 1.000) | 0.750 (0.726, 0.775) |
| Brainstem Ischemia | 9 | 0.817 (0.642, 1.000) | 1.000 (1.000, 1.000) | 0.222 (0.198, 0.246) |
| Empty Sella | 8 | 0.869 (0.743, 1.000) | 0.875 (0.750, 1.000) | 0.585 (0.556, 0.613) |
| Artery Tortuosity | 8 | 0.786 (0.604, 1.000) | 0.875 (0.750, 1.000) | 0.333 (0.306, 0.361) |
| Aneurysm | 8 | 0.822 (0.691, 0.979) | 0.625 (0.375, 1.000) | 1.000 (1.000, 1.000) |
| Ethmoid Fracture | 7 | 0.901 (0.815, 1.000) | 1.000 (1.000, 1.000) | 0.537 (0.507, 0.566) |
| Tuberous Sclerosis Complex | 6 | 0.806 (0.613, 1.000) | 0.833 (0.667, 1.000) | 0.699 (0.673, 0.726) |
| Gliomas | 5 | 0.999 (0.999, 1.000) | 1.000 (1.000, 1.000) | 0.997 (0.995, 1.000) |
| Subacute Cerebral Infarction | 5 | 0.872 (0.759, 1.000) | 1.000 (1.000, 1.000) | 0.525 (0.497, 0.553) |
| Insula Ischemia | 5 | 0.828 (0.684, 1.000) | 0.800 (0.600, 1.000) | 0.785 (0.761, 0.811) |
| Cerebellar Lesion | 4 | 0.887 (0.776, 1.000) | 1.000 (1.000, 1.000) | 0.624 (0.595, 0.651) |
| Ethmoid Paperboard Fracture | 4 | 0.815 (0.630, 1.000) | 0.750 (0.500, 1.000) | 0.743 (0.718, 0.767) |
| Pons Ischemia | 4 | 0.975 (0.950, 1.000) | 1.000 (1.000, 1.000) | 0.948 (0.935, 0.961) |
| Cerebral Fissure | 3 | 0.987 (0.973, 1.000) | 1.000 (1.000, 1.000) | 0.960 (0.948, 0.972) |
| Demyelination | 3 | 0.876 (0.752, 1.000) | 1.000 (1.000, 1.000) | 0.628 (0.601, 0.656) |
| Widened Ventricles | 3 | 0.975 (0.950, 1.000) | 1.000 (1.000, 1.000) | 0.926 (0.910, 0.941) |
| Widened Subdural Space | 3 | 1.000 (1.000, 1.000) | 1.000 (1.000, 1.000) | 1.000 (1.000, 1.000) |
| Cisterna Magna Cyst | 3 | 0.695 (0.389, 1.000) | 1.000 (1.000, 1.000) | 0.282 (0.256, 0.309) |
| Cerebral Hemisphere Compression | 2 | 1.000 (1.000, 1.000) | 1.000 (1.000, 1.000) | 1.000 (1.000, 1.000) |
| Bony Process | 2 | 0.625 (0.249, 1.000) | 1.000 (1.000, 1.000) | 0.249 (0.224, 0.274) |
| Arterial Thickening | 2 | 0.400 (0.229, 0.570) | 1.000 (1.000, 1.000) | 0.247 (0.223, 0.271) |
| Pineal Gland Enlargement | 2 | 0.951 (0.902, 1.000) | 1.000 (1.000, 1.000) | 0.902 (0.885, 0.920) |
| Dysplasia | 1 | 1.000 (1.000, 1.000) | 1.000 (1.000, 1.000) | 1.000 (1.000, 1.000) |
| Cerebellar Compression | 1 | 0.739 (0.712, 0.764) | 1.000 (1.000, 1.000) | 0.739 (0.712, 0.764) |
| Negative Occupancy Effect | 1 | 1.000 (1.000, 1.000) | 1.000 (1.000, 1.000) | 1.000 (1.000, 1.000) |
| Arachnoid Granules | 1 | 0.866 (0.847, 0.886) | 1.000 (1.000, 1.000) | 0.866 (0.847, 0.886) |
| Widened Tentorium | 1 | 1.000 (1.000, 1.000) | 1.000 (1.000, 1.000) | 1.000 (1.000, 1.000) |
| Widened Lambdoid Sutures | 1 | 0.932 (0.918, 0.948) | 1.000 (1.000, 1.000) | 0.932 (0.918, 0.948) |
| Ventricular Reduction | 1 | 1.000 (1.000, 1.000) | 1.000 (1.000, 1.000) | 1.000 (1.000, 1.000) |
| Adenoid Hypertrophy | 1 | 1.000 (1.000, 1.000) | 1.000 (1.000, 1.000) | 1.000 (1.000, 1.000) |
| Cystic Pineal Gland | 1 | 0.937 (0.924, 0.951) | 1.000 (1.000, 1.000) | 0.937 (0.924, 0.951) |
| Frontal Lobe Cyst | 1 | 1.000 (1.000, 1.000) | 1.000 (1.000, 1.000) | 1.000 (1.000, 1.000) |
| Neuroma | 1 | 0.997 (0.995, 1.000) | 1.000 (1.000, 1.000) | 0.997 (0.995, 1.000) |
| Thyroid Cartilage Fracture | 1 | 0.932 (0.917, 0.948) | 1.000 (1.000, 1.000) | 0.932 (0.917, 0.948) |
| Capsular Infarction | 1 | 1.000 (1.000, 1.000) | 1.000 (1.000, 1.000) | 1.000 (1.000, 1.000) |
| Massive Hemorrhage | 1 | 1.000 (1.000, 1.000) | 1.000 (1.000, 1.000) | 1.000 (1.000, 1.000) |
| avg | 127/127 | 0.883 (0.858, 0.911) | 0.810 (0.765, 0.858) | 0.835 (0.815, 0.856) |

**Supplementary Table 2. Performance on the prospective dataset for head disorder detection.**

| Prospective | | | | |
|---|---|---|---|---|
| | **Num** | **AUC** | **Sensitivity** | **Specificity** |
| Ischemia | 473 | 0.903 (0.884, 0.923) | 0.833 (0.801, 0.867) | 0.804 (0.761, 0.847) |
| Malacia Foci | 460 | 0.931 (0.916, 0.948) | 0.848 (0.817, 0.880) | 0.847 (0.807, 0.887) |
| Parenchymal Hemorrhage | 220 | 0.956 (0.941, 0.973) | 0.900 (0.864, 0.941) | 0.844 (0.801, 0.887) |
| Pneumocephaly | 213 | 0.931 (0.906, 0.958) | 0.878 (0.836, 0.925) | 0.847 (0.807, 0.887) |
| Subarachnoid Hemorrhage | 213 | 0.949 (0.928, 0.972) | 0.897 (0.859, 0.939) | 0.877 (0.844, 0.914) |
| Brainswelling | 193 | 0.830 (0.793, 0.868) | 0.777 (0.725, 0.839) | 0.715 (0.669, 0.761) |
| Effusion | 168 | 0.940 (0.918, 0.965) | 0.875 (0.827, 0.929) | 0.847 (0.807, 0.887) |
| Soft Tissue Swelling | 142 | 0.825 (0.782, 0.871) | 0.761 (0.690, 0.831) | 0.748 (0.702, 0.798) |
| Subdural Hematoma | 123 | 0.939 (0.911, 0.970) | 0.886 (0.837, 0.943) | 0.840 (0.801, 0.880) |
| Calcification | 115 | 0.792 (0.736, 0.850) | 0.757 (0.678, 0.835) | 0.718 (0.672, 0.767) |
| Lacunar Infarction | 109 | 0.662 (0.599, 0.729) | 0.596 (0.505, 0.688) | 0.684 (0.635, 0.733) |
| Contusion | 107 | 0.931 (0.902, 0.965) | 0.850 (0.785, 0.925) | 0.844 (0.807, 0.883) |
| Basal Ganglia Cerebral Infarction | 92 | 0.646 (0.577, 0.718) | 0.543 (0.446, 0.641) | 0.724 (0.675, 0.773) |
| Space Occupying Lesions | 90 | 0.880 (0.836, 0.927) | 0.800 (0.722, 0.878) | 0.748 (0.699, 0.798) |
| Intraventricular Hemorrhage | 83 | 0.956 (0.929, 0.990) | 0.904 (0.843, 0.964) | 0.847 (0.807, 0.883) |
| Sclerosis | 81 | 0.854 (0.799, 0.910) | 0.741 (0.642, 0.827) | 0.850 (0.810, 0.890) |
| Tubercle | 78 | 0.797 (0.735, 0.862) | 0.718 (0.628, 0.821) | 0.776 (0.730, 0.822) |
| Subcutaneous Hematoma | 78 | 0.829 (0.771, 0.892) | 0.782 (0.692, 0.885) | 0.755 (0.712, 0.804) |
| Midline Shift | 74 | 0.602 (0.519, 0.688) | 0.500 (0.392, 0.608) | 0.678 (0.629, 0.727) |
| Cavum Septum Pellucidum | 72 | 0.747 (0.680, 0.817) | 0.625 (0.514, 0.736) | 0.788 (0.745, 0.831) |
| Cerebral Infarction of Caudate Nucleus Head | 72 | 0.889 (0.841, 0.943) | 0.847 (0.764, 0.931) | 0.782 (0.736, 0.828) |
| Arachnoid Cyst | 67 | 0.658 (0.586, 0.738) | 0.642 (0.537, 0.761) | 0.604 (0.552, 0.656) |
| Cisterna Magna | 66 | 0.785 (0.720, 0.860) | 0.697 (0.591, 0.818) | 0.755 (0.712, 0.804) |
| Artifacts Shadow | 65 | 0.954 (0.924, 0.990) | 0.908 (0.846, 0.985) | 0.847 (0.810, 0.887) |
| Brain Atrophy | 63 | 0.963 (0.939, 0.996) | 0.921 (0.857, 1.000) | 0.893 (0.859, 0.926) |
| Epidural Hematoma | 62 | 0.958 (0.932, 0.990) | 0.903 (0.839, 0.984) | 0.868 (0.831, 0.905) |
| Basal Ganglia Ischemia | 61 | 0.821 (0.758, 0.887) | 0.738 (0.639, 0.852) | 0.752 (0.706, 0.798) |
| Minor Hemorrhage | 60 | 0.962 (0.937, 0.993) | 0.917 (0.850, 0.983) | 0.896 (0.862, 0.929) |
| Multiple Cerebral Infarctions | 59 | 0.836 (0.770, 0.910) | 0.763 (0.661, 0.881) | 0.807 (0.767, 0.850) |
| Cerebral Edema | 58 | 0.924 (0.881, 0.978) | 0.914 (0.845, 1.000) | 0.807 (0.764, 0.850) |
| Ventriculomegaly | 55 | 0.945 (0.912, 0.986) | 0.945 (0.891, 1.000) | 0.755 (0.709, 0.801) |
| Paraventricular Ischemia | 53 | 0.823 (0.763, 0.888) | 0.660 (0.547, 0.792) | 0.807 (0.767, 0.853) |
| Atherosclerosis | 48 | 0.930 (0.893, 0.973) | 0.875 (0.792, 0.979) | 0.847 (0.810, 0.887) |
| Scalp Hematoma | 48 | 0.797 (0.718, 0.878) | 0.667 (0.542, 0.792) | 0.837 (0.801, 0.877) |
| Periventricular Cerebral Infarction | 47 | 0.666 (0.577, 0.760) | 0.574 (0.447, 0.723) | 0.693 (0.644, 0.742) |
| Subdural Hemorrhage | 47 | 0.901 (0.856, 0.954) | 0.894 (0.809, 1.000) | 0.730 (0.684, 0.779) |
| Senile Encephalopathy | 44 | 0.978 (0.961, 1.000) | 0.977 (0.955, 1.000) | 0.896 (0.865, 0.929) |
| Hydrocephalus | 44 | 0.954 (0.916, 1.000) | 0.932 (0.864, 1.000) | 0.847 (0.810, 0.887) |
| Occipital Fracture | 42 | 0.823 (0.742, 0.910) | 0.810 (0.690, 0.929) | 0.736 (0.690, 0.785) |
| Osteoma | 41 | 0.789 (0.698, 0.887) | 0.732 (0.610, 0.854) | 0.755 (0.709, 0.804) |
| Parietal Fracture | 35 | 0.836 (0.756, 0.924) | 0.743 (0.600, 0.886) | 0.773 (0.727, 0.819) |
| Frontal Lobe Ischemia | 35 | 0.750 (0.662, 0.843) | 0.629 (0.457, 0.800) | 0.758 (0.712, 0.804) |

| Prospective | | | | |
|---|---|---|---|---|
| | **Num** | **AUC** | **Sensitivity** | **Specificity** |
| Parenchymal Hematoma | 35 | 0.961 (0.934, 1.000) | 0.914 (0.829, 1.000) | 0.847 (0.810, 0.887) |
| Herniation | 32 | 0.990 (0.979, 1.000) | 1.000 (1.000, 1.000) | 0.672 (0.620, 0.724) |
| Lipoma | 30 | 0.856 (0.788, 0.932) | 0.800 (0.667, 0.967) | 0.785 (0.742, 0.828) |
| Arteriosclerosis | 23 | 0.730 (0.601, 0.868) | 0.565 (0.348, 0.783) | 0.899 (0.868, 0.933) |
| Frontal Bone Fracture | 23 | 0.731 (0.613, 0.862) | 0.696 (0.522, 0.870) | 0.666 (0.617, 0.718) |
| Hemispheric Infarction | 23 | 0.991 (0.983, 1.000) | 0.957 (0.913, 1.000) | 0.951 (0.929, 0.975) |
| Absence Of Bone | 22 | 0.823 (0.729, 0.927) | 0.818 (0.682, 1.000) | 0.656 (0.604, 0.709) |
| Temporal Lobe Cerebral Infarction | 21 | 0.822 (0.709, 0.956) | 0.810 (0.667, 1.000) | 0.752 (0.706, 0.801) |
| Frontal Lobe Cerebral Infarction | 21 | 0.712 (0.551, 0.882) | 0.619 (0.429, 0.857) | 0.868 (0.831, 0.908) |
| Corona Radiata Cerebral Infarction | 18 | 0.614 (0.444, 0.794) | 0.556 (0.333, 0.778) | 0.752 (0.706, 0.801) |
| Parietal Lobe Ischemia | 17 | 0.827 (0.721, 0.953) | 0.824 (0.647, 1.000) | 0.669 (0.617, 0.718) |
| Occipital Lobe Cerebral Infarction | 16 | 0.943 (0.894, 1.000) | 0.875 (0.750, 1.000) | 0.804 (0.761, 0.850) |
| Zygomatic Fracture | 15 | 0.904 (0.830, 0.993) | 0.867 (0.733, 1.000) | 0.675 (0.623, 0.724) |
| Temporal Bone Fracture | 15 | 0.693 (0.511, 0.886) | 0.600 (0.333, 0.867) | 0.951 (0.929, 0.975) |
| Parietal Lobe Cerebral Infarction | 15 | 0.854 (0.736, 1.000) | 0.867 (0.733, 1.000) | 0.721 (0.675, 0.770) |
| Malformation | 14 | 0.800 (0.698, 0.910) | 0.786 (0.571, 1.000) | 0.727 (0.678, 0.776) |
| Widened Cisterna Magna | 14 | 0.872 (0.789, 0.965) | 0.857 (0.714, 1.000) | 0.758 (0.712, 0.804) |
| Cystic Foci | 14 | 0.675 (0.530, 0.827) | 0.571 (0.286, 0.857) | 0.718 (0.669, 0.767) |
| Wallerian Degeneration | 13 | 0.997 (0.994, 1.000) | 1.000 (1.000, 1.000) | 0.979 (0.963, 0.994) |
| Empty Sella | 13 | 0.931 (0.881, 1.000) | 0.923 (0.846, 1.000) | 0.816 (0.776, 0.856) |
| Septum Pellucidum Cyst | 13 | 0.515 (0.350, 0.696) | 0.462 (0.231, 0.769) | 0.764 (0.718, 0.810) |
| Skull Fracture | 13 | 0.900 (0.807, 1.000) | 0.923 (0.846, 1.000) | 0.733 (0.687, 0.779) |
| Metal Shadow | 12 | 0.923 (0.856, 1.000) | 0.917 (0.833, 1.000) | 0.899 (0.868, 0.933) |
| Zygomatic Arch Fracture | 12 | 0.895 (0.820, 0.993) | 0.833 (0.667, 1.000) | 0.788 (0.745, 0.834) |
| Centrum Semiovale Ischemia | 12 | 0.671 (0.492, 0.865) | 0.667 (0.417, 0.917) | 0.727 (0.678, 0.773) |
| Brainstem Ischemia | 12 | 0.841 (0.684, 1.000) | 0.833 (0.667, 1.000) | 0.604 (0.552, 0.660) |
| Osseous Defects | 11 | 0.886 (0.799, 1.000) | 0.909 (0.818, 1.000) | 0.623 (0.571, 0.672) |
| Widened Ventricles | 10 | 0.876 (0.752, 1.000) | 0.900 (0.800, 1.000) | 0.604 (0.549, 0.660) |
| Corona Radiata Ischemia | 10 | 0.728 (0.549, 0.921) | 0.700 (0.400, 1.000) | 0.629 (0.577, 0.678) |
| Cerebral Fissure | 8 | 0.729 (0.524, 0.987) | 0.625 (0.375, 1.000) | 0.963 (0.945, 0.985) |
| Fat Density | 8 | 0.777 (0.598, 1.000) | 0.875 (0.750, 1.000) | 0.531 (0.475, 0.586) |
| Insular Infarction | 8 | 0.855 (0.710, 1.000) | 0.875 (0.750, 1.000) | 0.537 (0.485, 0.595) |
| Cerebral White Matter Degeneration | 7 | 0.604 (0.331, 0.906) | 0.571 (0.286, 1.000) | 0.764 (0.718, 0.813) |
| Metastases | 7 | 0.970 (0.940, 1.000) | 1.000 (1.000, 1.000) | 0.868 (0.834, 0.905) |
| Thalamic Cerebral Infarction | 7 | 0.536 (0.332, 0.747) | 0.714 (0.429, 1.000) | 0.457 (0.402, 0.509) |
| Old Cerebral Infarction | 7 | 0.692 (0.457, 0.952) | 0.857 (0.714, 1.000) | 0.273 (0.224, 0.322) |
| Widened Subdural Space | 6 | 0.860 (0.728, 1.000) | 0.833 (0.667, 1.000) | 0.794 (0.752, 0.840) |
| Meningioma | 6 | 0.785 (0.601, 0.998) | 1.000 (1.000, 1.000) | 0.448 (0.393, 0.500) |
| Centrum Semiovale Cerebral Infarction | 6 | 0.522 (0.267, 0.787) | 0.667 (0.333, 1.000) | 0.503 (0.451, 0.555) |
| Bony Process | 5 | 0.434 (0.185, 0.675) | 0.400 (0.000, 0.800) | 0.733 (0.687, 0.779) |
| Fat Deposition | 5 | 0.958 (0.915, 1.000) | 1.000 (1.000, 1.000) | 0.788 (0.748, 0.831) |
| Tumour | 5 | 0.780 (0.577, 1.000) | 0.800 (0.600, 1.000) | 0.779 (0.733, 0.825) |

| Prospective | | | | |
|---|---|---|---|---|
| | **Num** | **AUC** | **Sensitivity** | **Specificity** |
| Multiple Hemorrhages | 5 | 1.000 (1.000, 1.000) | 1.000 (1.000, 1.000) | 1.000 (1.000, 1.000) |
| Enhancement Foci | 4 | 0.939 (0.879, 1.000) | 1.000 (1.000, 1.000) | 0.834 (0.794, 0.874) |
| Widened Tentorium | 4 | 1.000 (1.000, 1.000) | 1.000 (1.000, 1.000) | 1.000 (1.000, 1.000) |
| Cisterna Magna Cyst | 4 | 0.929 (0.859, 1.000) | 1.000 (1.000, 1.000) | 0.736 (0.690, 0.785) |
| Cerebellar Hemisphere Infarction | 4 | 0.781 (0.561, 1.000) | 1.000 (1.000, 1.000) | 0.334 (0.282, 0.383) |
| Arterial Thickening | 3 | 0.976 (0.953, 1.000) | 1.000 (1.000, 1.000) | 0.929 (0.905, 0.960) |
| Artery Tortuosity | 3 | 0.856 (0.764, 0.985) | 1.000 (1.000, 1.000) | 0.721 (0.675, 0.770) |
| Widened Arterial | 3 | 0.889 (0.777, 1.000) | 1.000 (1.000, 1.000) | 0.693 (0.638, 0.745) |
| Sphenoid Fracture | 3 | 0.942 (0.883, 1.000) | 1.000 (1.000, 1.000) | 0.825 (0.785, 0.868) |
| Brainstem Infarction | 3 | 0.940 (0.879, 1.000) | 1.000 (1.000, 1.000) | 0.868 (0.831, 0.908) |
| Subacute Cerebral Infarction | 3 | 0.951 (0.902, 1.000) | 1.000 (1.000, 1.000) | 0.902 (0.871, 0.933) |
| Epidural Hemorrhage | 3 | 0.984 (0.967, 1.000) | 1.000 (1.000, 1.000) | 0.951 (0.929, 0.975) |
| Brainstem Swelling | 2 | 0.992 (0.985, 1.000) | 1.000 (1.000, 1.000) | 0.985 (0.972, 1.000) |
| Arterial Cilia | 2 | 1.000 (1.000, 1.000) | 1.000 (1.000, 1.000) | 1.000 (1.000, 1.000) |
| Widened Subarachnoid Space | 2 | 0.739 (0.632, 0.850) | 1.000 (1.000, 1.000) | 0.660 (0.610, 0.712) |
| Myeloma | 2 | 0.502 (0.003, 1.000) | 1.000 (1.000, 1.000) | 0.003 (0.000, 0.003) |
| Dysplasia | 1 | 0.816 (0.776, 0.859) | 1.000 (1.000, 1.000) | 0.816 (0.776, 0.859) |
| Cerebral Hemisphere Compression | 1 | 0.991 (0.982, 1.000) | 1.000 (1.000, 1.000) | 0.991 (0.982, 1.000) |
| Arachnoid Granules | 1 | 0.985 (0.972, 1.000) | 1.000 (1.000, 1.000) | 0.985 (0.972, 1.000) |
| Leukoaraiosis | 1 | 0.816 (0.773, 0.859) | 1.000 (1.000, 1.000) | 0.816 (0.773, 0.859) |
| Cerebral Cortical Atrophy | 1 | 1.000 (1.000, 1.000) | 1.000 (1.000, 1.000) | 1.000 (1.000, 1.000) |
| Ventricular Reduction | 1 | 0.991 (0.982, 1.000) | 1.000 (1.000, 1.000) | 0.991 (0.982, 1.000) |
| Pineal Gland Enlargement | 1 | 0.991 (0.982, 1.000) | 1.000 (1.000, 1.000) | 0.991 (0.982, 1.000) |
| Sella Turcica Enlargement | 1 | 0.979 (0.963, 0.997) | 1.000 (1.000, 1.000) | 0.979 (0.963, 0.997) |
| Choroid Cyst | 1 | 0.629 (0.577, 0.681) | 1.000 (1.000, 1.000) | 0.629 (0.577, 0.681) |
| Frontal Lobe Cyst | 1 | 1.000 (1.000, 1.000) | 1.000 (1.000, 1.000) | 1.000 (1.000, 1.000) |
| Facial Bone Fracture | 1 | 0.258 (0.209, 0.301) | 1.000 (1.000, 1.000) | 0.258 (0.209, 0.301) |
| Pons Ischemia | 1 | 0.908 (0.877, 0.939) | 1.000 (1.000, 1.000) | 0.908 (0.877, 0.939) |
| Insula Ischemia | 1 | 0.736 (0.690, 0.785) | 1.000 (1.000, 1.000) | 0.736 (0.690, 0.785) |
| Minor Haematoma | 1 | 1.000 (1.000, 1.000) | 1.000 (1.000, 1.000) | 1.000 (1.000, 1.000) |
| Massive Hemorrhage | 1 | 1.000 (1.000, 1.000) | 1.000 (1.000, 1.000) | 1.000 (1.000, 1.000) |
| Cerebellar Vermis Hemorrhage | 1 | 0.828 (0.788, 0.868) | 1.000 (1.000, 1.000) | 0.828 (0.788, 0.868) |
| avg | 116/116 | 0.868 (0.821, 0.918) | 0.809 (0.737, 0.888) | 0.795 (0.753, 0.838) |

**Supplementary Table 3. Performance on the cross-center dataset for head disorder detection.**

| Cross-center | | | | |
|---|---|---|---|---|
| | Num | AUC | Sensitivity | Specificity |
| Lacunar Infarction | 100 | 0.862 (0.861, 0.864) | 0.839 (0.837, 0.842) | 0.781 (0.779, 0.784) |
| Senile Encephalopathy | 99 | 0.879 (0.877, 0.880) | 0.848 (0.845, 0.852) | 0.788 (0.784, 0.791) |
| Subarachnoid Hemorrhage | 96 | 0.903 (0.901, 0.904) | 0.818 (0.815, 0.821) | 0.869 (0.866, 0.872) |
| Parenchymal Hemorrhage | 88 | 0.843 (0.840, 0.844) | 0.778 (0.775, 0.781) | 0.796 (0.793, 0.798) |
| Contusion | 64 | 0.858 (0.854, 0.857) | 0.731 (0.728, 0.734) | 0.877 (0.874, 0.880) |
| Malacia Foci | 56 | 0.843 (0.842, 0.846) | 0.784 (0.781, 0.788) | 0.809 (0.806, 0.812) |
| Subdural Hematoma | 52 | 0.877 (0.875, 0.879) | 0.834 (0.831, 0.837) | 0.857 (0.854, 0.861) |
| Parenchymal Hematoma | 43 | 0.798 (0.796, 0.801) | 0.677 (0.674, 0.680) | 0.854 (0.851, 0.857) |
| Intraventricular Hemorrhage | 41 | 0.923 (0.922, 0.925) | 0.855 (0.853, 0.857) | 0.968 (0.966, 0.969) |
| Soft Tissue Injury | 39 | 0.857 (0.855, 0.858) | 0.719 (0.716, 0.722) | 0.966 (0.965, 0.968) |
| Epidural Hematoma | 22 | 0.872 (0.869, 0.873) | 0.772 (0.769, 0.775) | 1.000 (1.000, 1.000) |
| Leukoaraiosis | 21 | 0.870 (0.867, 0.870) | 0.821 (0.816, 0.826) | 0.773 (0.769, 0.778) |
| Brain Atrophy | 19 | 0.884 (0.881, 0.884) | 0.789 (0.786, 0.791) | 0.932 (0.930, 0.934) |
| Pneumocephaly | 19 | 0.941 (0.941, 0.943) | 0.895 (0.893, 0.897) | 0.942 (0.941, 0.944) |
| Effusion | 17 | 0.914 (0.913, 0.916) | 0.904 (0.901, 0.906) | 0.910 (0.907, 0.912) |
| Scalp Hematoma | 14 | 0.971 (0.970, 0.972) | 0.928 (0.926, 0.929) | 1.000 (1.000, 1.000) |
| Brainswelling | 12 | 0.924 (0.921, 0.923) | 0.831 (0.829, 0.833) | 1.000 (1.000, 1.000) |
| Herniation | 9 | 0.833 (0.832, 0.835) | 0.758 (0.754, 0.763) | 0.797 (0.792, 0.802) |
| Soft Tissue Swelling | 7 | 0.952 (0.952, 0.954) | 0.859 (0.857, 0.861) | 1.000 (0.999, 1.000) |
| Occipital Lobe Cerebral Infarction | 6 | 0.944 (0.943, 0.945) | 0.833 (0.831, 0.836) | 1.000 (1.000, 1.000) |
| Basal Ganglia Cerebral Infarction | 5 | 1.000 (1.000, 1.000) | 1.000 (1.000, 1.000) | 1.000 (1.000, 1.000) |
| Arachnoid Cyst | 4 | 0.625 (0.622, 0.628) | 0.530 (0.525, 0.535) | 0.956 (0.948, 0.965) |
| Frontal Lobe Cerebral Infarction | 4 | 0.625 (0.624, 0.629) | 0.634 (0.627, 0.642) | 0.640 (0.632, 0.647) |
| Calcification | 3 | 1.000 (1.000, 1.000) | 1.000 (1.000, 1.000) | 1.000 (1.000, 1.000) |
| Parietal Lobe Cerebral Infarction | 3 | 0.667 (0.661, 0.667) | 1.000 (1.000, 1.000) | 0.664 (0.661, 0.667) |
| Cerebellar Hemisphere Infarction | 3 | 0.778 (0.774, 0.779) | 0.665 (0.662, 0.668) | 1.000 (1.000, 1.000) |
| Temporal Lobe Cerebral Infarction | 3 | 0.778 (0.776, 0.780) | 0.666 (0.663, 0.669) | 1.000 (1.000, 1.000) |
| Wallerian Degeneration | 3 | 0.889 (0.887, 0.890) | 0.841 (0.831, 0.851) | 0.853 (0.843, 0.863) |
| Insular Infarction | 3 | 0.778 (0.775, 0.779) | 1.000 (1.000, 1.000) | 0.666 (0.663, 0.669) |
| Hemispheric Infarction | 2 | 1.000 (1.000, 1.000) | 1.000 (1.000, 1.000) | 1.000 (1.000, 1.000) |
| Cerebral Edema | 2 | 1.000 (1.000, 1.000) | 1.000 (1.000, 1.000) | 1.000 (1.000, 1.000) |
| Subdural Hemorrhage | 1 | 1.000 (1.000, 1.000) | 1.000 (1.000, 1.000) | 1.000 (1.000, 1.000) |
| Multiple Hemorrhages | 1 | 0.000 (0.000, 0.000) | 0.000 (0.000, 0.000) | 0.000 (0.000, 0.000) |
| Subcutaneous Hematoma | 1 | 1.000 (1.000, 1.000) | 1.000 (1.000, 1.000) | 1.000 (1.000, 1.000) |
| Minor Hemorrhage | 1 | 0.000 (0.000, 0.000) | 0.000 (0.000, 0.000) | 0.000 (0.000, 0.000) |
| Brainstem Infarction | 1 | 0.000 (0.000, 0.000) | 0.000 (0.000, 0.000) | 0.000 (0.000, 0.000) |
| Temporal Bone Fracture | 1 | 1.000 (1.000, 1.000) | 1.000 (1.000, 1.000) | 1.000 (1.000, 1.000) |
| Fat Density | 1 | 1.000 (1.000, 1.000) | 1.000 (1.000, 1.000) | 1.000 (1.000, 1.000) |
| Midline Shift | 1 | 1.000 (1.000, 1.000) | 1.000 (1.000, 1.000) | 1.000 (1.000, 1.000) |
| Thalamic Cerebral Infarction | 1 | 0.000 (0.000, 0.000) | 0.000 (0.000, 0.000) | 0.000 (0.000, 0.000) |
| Cerebellar Vermis Hemorrhage | 1 | 0.000 (0.000, 0.000) | 0.000 (0.000, 0.000) | 0.000 (0.000, 0.000) |
| Subacute Cerebral Infarction | 1 | 1.000 (1.000, 1.000) | 1.000 (1.000, 1.000) | 1.000 (1.000, 1.000) |
| Malformation | 1 | 1.000 (1.000, 1.000) | 1.000 (1.000, 1.000) | 1.000 (1.000, 1.000) |
| Hydrocephalus | 1 | 0.000 (0.000, 0.000) | 0.000 (0.000, 0.000) | 0.000 (0.000, 0.000) |
| Centrum Semiovale Cerebral Infarction | 1 | 1.000 (1.000, 1.000) | 1.000 (1.000, 1.000) | 1.000 (1.000, 1.000) |
| Periventricular Cerebral Infarction | 1 | 1.000 (1.000, 1.000) | 1.000 (1.000, 1.000) | 1.000 (1.000, 1.000) |
| avg | 46/46 | 0.866 (0.865, 0.868) | 0.803 (0.800, 0.806) | 0.852 (0.850, 0.855) |

**Supplementary Table 4. True positive rate (TPR) at different false positive rate (FPR) levels.** This table presents the performance of the model at various FPR levels on the retrospective, prospective, and cross-center datasets. The TPR values were obtained by adjusting the decision threshold of the model, providing a comprehensive view of the model's performance across different levels of specificity.

| | | **True positive ratio (TPR)** | | |
|---|---|---|---|---|
| | | **Retrospective** | **Prospective** | **Cross-Center** |
| **False positive ratio (FPR)** | 0.1 | 0.651 | 0.588 | 0.606 |
| | 0.2 | 0.857 | 0.834 | 0.817 |
| | 0.3 | 0.891 | 0.875 | 0.859 |
| | 0.4 | 0.926 | 0.917 | 0.914 |
| | 0.5 | 0.955 | 0.955 | 0.954 |
| | 0.6 | 0.977 | 0.977 | 0.969 |
| | 0.7 | 1.000 | 1.000 | 1.000 |
| | 0.8 | 1.000 | 1.000 | 1.000 |
| | 0.9 | 1.000 | 1.000 | 1.000 |
| | 1.0 | 1.000 | 1.000 | 1.000 |

**Supplementary Table 5. Data characteristics of healthy scans in the training dataset, as well as those in the prospective, retrospective, and cross-center test datasets.** Scans in the training dataset (21,429) and the healthy Scans in the retrospective test dataset (1,173 scans) are from Mar. 2012 to Jul. 2019 (from the PLAGH). healthy scans in the prospective test dataset (345 scans) are from Jul. 2019 to Aug. 2021 (from the PLAGH). Healthy cans in the cross-center test dataset (227 scans) are from Apr. 2018 to May. 2019 (from the BHHP). The data characteristics encompass patient gender, age, and inspection time statistics.

| | **Gender** | | **Age** | | | **Inspection time** | | | **Total** |
|---|---|---|---|---|---|---|---|---|---|
| | **Male** | **Female** | **~44** | **44~59** | **59~** | **~2014** | **2014~2016** | **2016~2019** | **-** |
| **Training** | 13,227 | 8,202 | 7,036 | 11,333 | 3,060 | 2,030 | 10,516 | 8,883 | 21,429 |
| **Retrospective** | 645 | 528 | 428 | 527 | 218 | 145 | 602 | 426 | 1,173 |
| | **Male** | **Female** | **~44** | **44~59** | **59~** | **2019~2020** | **2020~** | | |
| **Prospective** | 190 | 155 | 155 | 136 | 54 | 135 | 210 | | 345 |
| | **Male** | **Female** | **~44** | **44~59** | **59~** | **2018~2019** | **2019~** | | |
| **Cross-center** | 96 | 131 | 134 | 71 | 22 | 65 | 162 | | 227 |

**Supplementary Table 6. Proportion of disorder and non-disorder samples in the prospective, retrospective, and cross-center test datasets.**

| | **Disorder** | **Non-Disorder** |
|---|---|---|
| **Retrospective** | 8,794 (88.2%) | 1,173 (11.8%) |
| **Prospective** | 2,709 (88.7%) | 345 (11.3%) |
| **Cross-center** | 327 (59.0%) | 227 (41.0%) |

**Supplementary Table 7. Classification of Disorder Types Across Different Lesion Sizes.** This table presents the classification of disorder types across different lesion sizes, categorized as large, medium, small, and other. The ’other’ category represents diseases with significant variations in lesion size.

| Size level | Disorder types |
| --- | --- |
| Large (19.7%) | 1. Brainswelling, 2. Cerebral Edema, 3. Cerebral White Matter Degeneration, 4. Cerebral Hemisphere Compression, 5. Cerebellar Compression, 6. Demyelination, 7. Epidural Hematoma, 8. Epidural Hemorrhage, 9. Gliomas, 10. Hydrocephalus, 11. Herniation, 12. Multiple Hemorrhages, 13. Midline Shift, 14. Meningioma, 15. Massive Hemorrhage, 16. Parenchymal Hemorrhage, 17. Subarachnoid Hemorrhage, 18. Subdural Hematoma, 19. Subdural Hemorrhage, 20. Tubercle, 21. Tumour, 22. Ventriculomegaly, 23. Widened Cisterna Magna, 24. Widened Subarachnoid Space, 25. Widened Ventricles |
| Medium (16.5%) | 1. Arachnoid Cyst, 2. Contusion, 3. Effusion, 4. Ethmoid Fracture, 5. Ethmoid Paperboard Fracture, 6. Frontal Bone Fracture, 7. Facial Bone Fracture, 8. Intraventricular Hemorrhage, 9. Leukoaraiosis, 10. Occipital Fracture, 11. Parenchymal Hematoma, 12. Parietal Fracture, 13. Skull Fracture, 14. Sphenoid Fracture, 15. Senile Encephalopathy, 16. Skull Base Fracture, 17. Temporal Bone Fracture, 18. Widened Subdural Space, 19. Widened Tentorium, 20. Zygomatic Fracture, 21. Zygomatic Arch Fracture |
| Small (32.3%) | 1. Arteriosclerosis, 2. Artifacts Shadow, 3. Atherosclerosis, 4. Artery Tortuosity, 5. Arterial Thickening, 6. Arachnoid Granules, 7. Adenoid Hypertrophy, 8. Bony Process, 9. Calcification, 10. Cavum Septum Pellucidum, 11. Cystic Foci, 12. Choroid Cyst, 13. Cerebral Fissure, 14. Cisterna Magna Cyst, 15. Cystic Pineal Gland, 16. Dysplasia, 17. Empty Sella, 18. Fat Deposition, 19. Fat Density, 20. Frontal Lobe Cyst, 21. Lipoma, 22. Malacia Foci, 23. Minor Hemorrhage, 24. Metal Shadow, 25. Minor Haematoma, 26. Malformation, 27. Negative Occupancy Effect, 28. Neuroma, 29. Osteoma, 30. Osseous Defects, 31. Pineal Gland Enlargement, 32. Sclerosis, 33. Soft Tissue Swelling, 34. Subcutaneous Hematoma, 35. Scalp Hematoma, 36. Sella Turcica Enlargement, 37. Septum Pellucidum Cyst, 38. Thyroid Cartilage Fracture, 39. Ventricular Reduction, 40. Wallerian Degeneration, 41. Widened Lambdoid Sutures |
| Others (31.5%) | 1. Absence Of Bone, 2. Aneurysm, 3. Basal Ganglia Ischemia, 4. Basal Ganglia Cerebral Infarction, 5. Brain Atrophy, 6. Brainstem Infarction, 7. Brainstem Ischemia, 8. Corona Radiata Cerebral Infarction, 9. Cerebral Infarction of Caudate Nucleus Head, 10. Centrum Semiovale Cerebral Infarction, 11. Cisterna Magna, 12. Centrum Semiovale Ischemia, 13. Corona Radiata Ischemia, 14. Cerebellar Hemisphere Infarction, 15. Cerebellar Lesion, 16. Capsular Infarction, 17. Enhancement Foci, 18. Frontal Lobe Cerebral Infarction, 19. Frontal Lobe Ischemia, 20. Hemispheric Infarction, 21. Ischemia, 22. Insular Infarction, 23. Insula Ischemia, 24. Lacunar Infarction, 25. Lacunar Ischemia, 26. Multiple Cerebral Infarctions, 27. Metastases, 28. Occipital Lobe Cerebral Infarction, 29. Old Cerebral Infarction, 30. Pneumocephaly, 31. Periventricular Cerebral Infarction, 32. Parietal Lobe Cerebral Infarction, 33. Paraventricular Ischemia, 34. Parietal Lobe Ischemia, 35. Pons Ischemia, 36. Space Occupying Lesions, 37. Subacute Cerebral Infarction, 38. Thalamic Cerebral Infarction, 39. Temporal Lobe Cerebral Infarction, 40. Tuberous Sclerosis Complex |

**Supplementary Table 8. Classification of disorder types based on urgency of treatment.** This table presents the classification of disorder types based on urgency of treatment, categorized as high, medium, and low urgency. The 'high urgency' category represents diseases requiring emergency intervention, such as certain types of cancer and other potentially life-threatening conditions. The 'medium urgency' category includes diseases that may not require immediate treatment but may need intervention as they progress over time. The 'low urgency' category includes diseases that typically do not require treatment intervention and have a minimal impact on the patient's quality of life.

| Intervention level | Disorder types |
|---|---|
| Emergency intervention (22.0%) | 1. Contusion, 2. Cerebral Infarction Of Caudate Nucleus Head, 3. Cerebellar Compression, 4. Effusion, 5. Epidural Hematoma, 6. Epidural Hemorrhage, 7. Ethmoid Fracture, 8. Ethmoid Paperboard Fracture, 9. Frontal Bone Fracture, 10. Gliomas, 11. Herniation, 12. Ischemia, 13. Massive Hemorrhage, 14. Occipital Fracture, 15. Parenchymal Hemorrhage, 16. Parietal Fracture, 17. Subarachnoid Hemorrhage, 18. Subdural Hematoma, 19. Subdural Hemorrhage, 20. Skull Fracture, 21. Sphenoid Fracture, 22. Skull Base Fracture, 23. Subacute Cerebral Infarction, 24. Temporal Bone Fracture, 25. Thyroid Cartilage Fracture, 26. Ventriculomegaly, 27. Zygomatic Fracture, 28. Zygomatic Arch Fracture |
| Selective intervention (30.0%) | 1. Arteriosclerosis, 2. Absence Of Bone, 3. Atherosclerosis, 4. Aneurysm, 5. Basal Ganglia Cerebral Infarction, 6. Brainswelling, 7. Brainstem Infarction, 8. Corona Radiata Cerebral Infarction, 9. Centrum Semiovale Cerebral Infarction, 10. Cerebellar Hemisphere Infarction, 11. Cerebellar Lesion, 12. Cerebral Hemisphere Compression, 13. Capsular Infarction, 14. Frontal Lobe Cerebral Infarction, 15. Hydrocephalus, 16. Hemispheric Infarction, 17. Intraventricular Hemorrhage, 18. Insular Infarction, 19. Lipoma, 20. Multiple Cerebral Infarctions, 21. Minor Hemorrhage, 22. Metastases, 23. Meningioma, 24. Negative Occupancy Effect, 25. Neuroma, 26. Occipital Lobe Cerebral Infarction, 27. Osteoma, 28. Old Cerebral Infarction, 29. Osseous Defects, 30. Pneumocephaly, 31. Periventricular Cerebral Infarction, 32. Parietal Lobe Cerebral Infarction, 33. Sclerosis, 34. Space Occupying Lesions, 35. Soft Tissue Swelling, 36. Thalamic Cerebral Infarction, 37. Temporal Lobe Cerebral Infarction, 38. Tumour |
| Non-intervention (48.0%) | 1. Arachnoid Cyst, 2. Artifacts Shadow, 3. Artery Tortuosity, 4. Arterial Thickening, 5. Arachnoid Granules, 6. Adenoid Hypertrophy, 7. Basal Ganglia Ischemia, 8. Brain Atrophy, 9. Brainstem Ischemia, 10. Bony Process, 11. Calcification, 12. Cerebral Edema, 13. Cerebral White Matter Degeneration, 14. Cavum Septum Pellucidum, 15. Cisterna Magna, 16. Centrum Semiovale Ischemia, 17. Corona Radiata Ischemia, 18. Cystic Foci, 19. Choroid Cyst, 20. Cerebral Fissure, 21. Cisterna Magna Cyst, 22. Cystic Pineal Gland, 23. Demyelination, 24. Dysplasia, 25. Enhancement Foci, 26. Empty Sella, 27. Frontal Lobe Ischemia, 28. Facial Bone Fracture, 29. Fat Deposition, 30. Fat Density, 31. Frontal Lobe Cyst, 32. Insula Ischemia, 33. Lacunar Infarction, 34. Leukoaraiosis, 35. Lacunar Ischemia, 36. Malacia Foci, 37. Multiple Hemorrhages, 38. Metal Shadow, 39. Midline Shift, 40. Minor Haematoma, 41. Malformation, 42. Parenchymal Hematoma, 43. Paraventricular Ischemia, 44. Parietal Lobe Ischemia, 45. Pons Ischemia, 46. Pineal Gland Enlargement, 47. Subcutaneous Hematoma, 48. Scalp Hematoma, 49. Sella Turcica Enlargement, 50. Septum Pellucidum Cyst, 51. Senile Encephalopathy, 52. Tubercle, 53. Tuberous Sclerosis Complex, 54. Ventricular Reduction, 55. Widened Cisterna Magna, 56. Widened Subarachnoid Space, 57. Wallerian Degeneration, 58. Widened Ventricles, 59. Widened Subdural Space, 60. Widened Tentorium, 61. Widened Lambdoid Sutures |

**Supplementary Table 9. Statistics of the disorder-containing scans in the retrospective dataset.** The retrospective dataset comprises 8,794 scans with 127 types of disorders. These scans were obtained from the PLAGH and span from Mar. 2012 to Jul. 2019. Noticed that some scans exhibit multiple types of disorders, meaning that the total number of disorders is not equal to the sum of individual disorders.

| Retrospective | | | | | | | | | |
|---|---|---|---|---|---|---|---|---|---|
| | **Gender** | | **Age** | | | **Inspection time** | | | **Total** |
| | **Male** | **Female** | **~44** | **44~59** | **59~** | **~2014** | **2014~2016** | **2016~2019** | **-** |
| Effusion | 329 | 158 | 82 | 88 | 317 | 41 | 213 | 233 | 487 |
| Subarachnoid Hemorrhage | 289 | 196 | 91 | 152 | 242 | 46 | 241 | 198 | 485 |
| Subdural Hematoma | 344 | 138 | 113 | 92 | 277 | 44 | 250 | 188 | 482 |
| Pneumocephaly | 291 | 183 | 140 | 164 | 170 | 27 | 145 | 302 | 474 |
| Parenchymal Hemorrhage | 319 | 155 | 94 | 134 | 246 | 71 | 220 | 183 | 474 |
| Multiple Cerebral Infarctions | 304 | 161 | 9 | 91 | 365 | 115 | 206 | 144 | 465 |
| Corona Radiata Cerebral Infarction | 329 | 130 | 16 | 167 | 276 | 42 | 245 | 172 | 459 |
| Lacunar Infarction | 301 | 155 | 10 | 138 | 308 | 62 | 237 | 157 | 456 |
| Basal Ganglia Ischemia | 296 | 158 | 30 | 120 | 304 | 82 | 262 | 110 | 454 |
| Basal Ganglia Cerebral Infarction | 298 | 155 | 12 | 140 | 301 | 76 | 234 | 143 | 453 |
| Ischemia | 274 | 170 | 8 | 73 | 363 | 27 | 216 | 201 | 444 |
| Calcification | 229 | 213 | 79 | 131 | 232 | 34 | 229 | 179 | 442 |
| Malacia Foci | 312 | 124 | 40 | 104 | 292 | 34 | 215 | 187 | 436 |
| Contusion | 329 | 95 | 155 | 113 | 156 | 44 | 232 | 148 | 424 |
| Cerebral Infarction Of Caudate Nucleus Head | 294 | 128 | 21 | 150 | 251 | 53 | 148 | 221 | 422 |
| Periventricular Cerebral Infarction | 262 | 135 | 16 | 90 | 291 | 35 | 175 | 187 | 397 |
| Tubercle | 203 | 159 | 88 | 128 | 146 | 29 | 205 | 128 | 362 |
| Intraventricular Hemorrhage | 198 | 125 | 58 | 78 | 187 | 49 | 158 | 116 | 323 |
| Brainswelling | 214 | 101 | 101 | 102 | 112 | 24 | 144 | 147 | 315 |
| Sclerosis | 175 | 97 | 42 | 32 | 198 | 1 | 169 | 102 | 272 |
| Space Occupying Lesions | 141 | 120 | 42 | 90 | 129 | 46 | 131 | 84 | 261 |
| Epidural Hematoma | 185 | 67 | 123 | 78 | 51 | 49 | 114 | 89 | 252 |
| Cerebral Edema | 136 | 80 | 44 | 70 | 102 | 20 | 111 | 85 | 216 |
| Minor Hemorrhage | 104 | 108 | 63 | 74 | 75 | 29 | 74 | 109 | 212 |
| Thalamic Cerebral Infarction | 153 | 52 | 5 | 71 | 129 | 27 | 117 | 61 | 205 |
| Soft Tissue Swelling | 148 | 55 | 71 | 58 | 74 | 8 | 85 | 110 | 203 |
| Arteriosclerosis | 127 | 69 | 0 | 31 | 165 | 1 | 143 | 52 | 196 |
| Parenchymal Hematoma | 118 | 58 | 43 | 45 | 88 | 33 | 85 | 58 | 176 |
| Centrum Semiovale Cerebral Infarction | 93 | 58 | 4 | 51 | 96 | 19 | 90 | 42 | 151 |
| Parietal Lobe Cerebral Infarction | 97 | 51 | 10 | 63 | 75 | 15 | 89 | 44 | 148 |
| Frontal Lobe Cerebral Infarction | 91 | 32 | 6 | 44 | 73 | 18 | 76 | 29 | 123 |
| Arachnoid Cyst | 84 | 35 | 49 | 24 | 46 | 6 | 55 | 58 | 119 |
| Hydrocephalus | 78 | 30 | 41 | 25 | 42 | 7 | 46 | 55 | 108 |
| Cerebral White Matter Degeneration | 68 | 39 | 1 | 6 | 100 | 10 | 65 | 32 | 107 |
| Paraventricular Ischemia | 65 | 39 | 5 | 34 | 65 | 12 | 60 | 32 | 104 |
| Cavum Septum Pellucidum | 68 | 34 | 27 | 31 | 44 | 2 | 40 | 60 | 102 |
| Subcutaneous Hematoma | 66 | 36 | 35 | 23 | 44 | 3 | 62 | 37 | 102 |
| Temporal Bone Fracture | 81 | 20 | 63 | 29 | 9 | 17 | 63 | 21 | 101 |
| Frontal Lobe Ischemia | 56 | 36 | 2 | 44 | 46 | 13 | 43 | 36 | 92 |
| Subdural Hemorrhage | 62 | 27 | 19 | 14 | 56 | 9 | 37 | 43 | 89 |
| Ventriculomegaly | 51 | 32 | 32 | 16 | 35 | 5 | 32 | 46 | 83 |

| Retrospective | | | | | | | | | |
|---|---|---|---|---|---|---|---|---|---|
| | **Gender** | | **Age** | | | **Inspection time** | | | **Total** |
| | **Male** | **Female** | **~44** | **44~59** | **59~** | **~2014** | **2014~2016** | **2016~2019** | **-** |
| Parietal Fracture | 60 | 23 | 38 | 27 | 18 | 7 | 51 | 25 | 83 |
| Occipital Fracture | 66 | 16 | 41 | 16 | 25 | 15 | 41 | 26 | 82 |
| Occipital Lobe Cerebral Infarction | 49 | 24 | 10 | 28 | 35 | 8 | 41 | 24 | 73 |
| Frontal Bone Fracture | 65 | 6 | 33 | 18 | 20 | 4 | 43 | 24 | 71 |
| Osteoma | 31 | 39 | 4 | 18 | 48 | 2 | 24 | 44 | 70 |
| Temporal Lobe Cerebral Infarction | 47 | 19 | 7 | 33 | 26 | 7 | 37 | 22 | 66 |
| Old Cerebral Infarction | 49 | 17 | 2 | 30 | 34 | 16 | 35 | 15 | 66 |
| Lipoma | 45 | 18 | 5 | 7 | 51 | 6 | 27 | 30 | 63 |
| Scalp Hematoma | 33 | 30 | 33 | 11 | 19 | 8 | 36 | 19 | 63 |
| Artifacts Shadow | 31 | 31 | 10 | 19 | 33 | 3 | 36 | 23 | 62 |
| Brain Atrophy | 39 | 20 | 7 | 12 | 40 | 7 | 27 | 25 | 59 |
| Cisterna Magna | 49 | 10 | 14 | 22 | 23 | 4 | 17 | 38 | 59 |
| Herniation | 30 | 25 | 15 | 12 | 28 | 3 | 33 | 19 | 55 |
| Skull Fracture | 51 | 3 | 28 | 19 | 7 | 1 | 39 | 14 | 54 |
| Absence Of Bone | 37 | 13 | 13 | 11 | 26 | 6 | 19 | 25 | 50 |
| Hemispheric Infarction | 41 | 9 | 2 | 5 | 43 | 5 | 36 | 9 | 50 |
| Brainstem Infarction | 39 | 9 | 5 | 23 | 20 | 1 | 36 | 11 | 48 |
| Multiple Hemorrhages | 29 | 18 | 11 | 14 | 22 | 11 | 20 | 16 | 47 |
| Metal Shadow | 17 | 23 | 9 | 15 | 16 | 8 | 23 | 9 | 40 |
| Metastases | 28 | 12 | 2 | 13 | 25 | 6 | 26 | 8 | 40 |
| Centrum Semiovale Ischemia | 21 | 18 | 3 | 15 | 21 | 6 | 13 | 20 | 39 |
| Atherosclerosis | 24 | 14 | 1 | 1 | 36 | 1 | 6 | 31 | 38 |
| Midline Shift | 16 | 21 | 11 | 12 | 14 | 7 | 12 | 18 | 37 |
| Facial Bone Fracture | 23 | 8 | 19 | 11 | 1 | 0 | 13 | 18 | 31 |
| Corona Radiata Ischemia | 22 | 9 | 1 | 8 | 22 | 2 | 17 | 12 | 31 |
| Leukoaraiosis | 10 | 16 | 0 | 0 | 26 | 4 | 19 | 3 | 26 |
| Insular Infarction | 13 | 13 | 0 | 11 | 15 | 0 | 11 | 15 | 26 |
| Parietal Lobe Ischemia | 16 | 6 | 0 | 11 | 11 | 4 | 5 | 13 | 22 |
| Cerebellar Hemisphere Infarction | 12 | 9 | 5 | 9 | 7 | 1 | 17 | 3 | 21 |
| Minor Haematoma | 10 | 11 | 6 | 9 | 6 | 0 | 10 | 11 | 21 |
| Osseous Defects | 18 | 2 | 2 | 4 | 14 | 2 | 14 | 4 | 20 |
| Lacunar Ischemia | 16 | 3 | 1 | 1 | 17 | 4 | 11 | 4 | 19 |
| Zygomatic Fracture | 15 | 2 | 3 | 9 | 5 | 4 | 7 | 6 | 17 |
| Widened Cisterna Magna | 9 | 7 | 8 | 2 | 6 | 0 | 7 | 9 | 16 |
| Meningioma | 10 | 6 | 1 | 9 | 6 | 1 | 13 | 2 | 16 |
| Tumour | 9 | 7 | 5 | 6 | 5 | 1 | 10 | 5 | 16 |
| Widened Subarachnoid Space | 12 | 3 | 2 | 4 | 9 | 0 | 9 | 6 | 15 |
| Sphenoid Fracture | 12 | 2 | 14 | 0 | 0 | 8 | 1 | 5 | 14 |
| Malformation | 4 | 9 | 7 | 5 | 1 | 4 | 7 | 2 | 13 |
| Cystic Foci | 8 | 5 | 7 | 3 | 3 | 2 | 4 | 7 | 13 |
| Sella Turcica Enlargement | 5 | 7 | 2 | 4 | 6 | 0 | 8 | 4 | 12 |
| Choroid Cyst | 11 | 1 | 1 | 5 | 6 | 0 | 4 | 8 | 12 |
| Septum Pellucidum Cyst | 6 | 6 | 1 | 4 | 7 | 1 | 4 | 7 | 12 |

| Retrospective | | | | | | | | | |
|---|---|---|---|---|---|---|---|---|---|
| | Gender | | Age | | | Inspection time | | | Total |
| | Male | Female | ~44 | 44~59 | 59~ | ~2014 | 2014~2016 | 2016~2019 | - |
| Wallerian Degeneration | 9 | 2 | 0 | 3 | 8 | 1 | 5 | 5 | 11 |
| Senile Encephalopathy | 7 | 4 | 0 | 0 | 11 | 1 | 7 | 3 | 11 |
| Enhancement Foci | 10 | 1 | 1 | 3 | 7 | 0 | 3 | 8 | 11 |
| Skull Base Fracture | 11 | 0 | 3 | 4 | 4 | 4 | 6 | 1 | 11 |
| Zygomatic Arch Fracture | 10 | 1 | 2 | 6 | 3 | 3 | 5 | 3 | 11 |
| Epidural Hemorrhage | 7 | 3 | 1 | 6 | 3 | 0 | 7 | 3 | 10 |
| Fat Deposition | 2 | 7 | 0 | 1 | 8 | 0 | 8 | 1 | 9 |
| Fat Density | 2 | 7 | 2 | 2 | 5 | 0 | 7 | 2 | 9 |
| Brainstem Ischemia | 8 | 1 | 1 | 7 | 1 | 3 | 3 | 3 | 9 |
| Empty Sella | 1 | 7 | 0 | 4 | 4 | 1 | 6 | 1 | 8 |
| Artery Tortuosity | 6 | 2 | 0 | 5 | 3 | 2 | 2 | 4 | 8 |
| Aneurysm | 1 | 7 | 1 | 4 | 3 | 2 | 3 | 3 | 8 |
| Ethmoid Fracture | 7 | 0 | 3 | 2 | 2 | 1 | 2 | 4 | 7 |
| Tuberous Sclerosis Complex | 5 | 1 | 6 | 0 | 0 | 0 | 2 | 4 | 6 |
| Gliomas | 3 | 2 | 3 | 2 | 0 | 2 | 2 | 1 | 5 |
| Subacute Cerebral Infarction | 3 | 2 | 1 | 2 | 2 | 0 | 5 | 0 | 5 |
| Insula Ischemia | 2 | 3 | 0 | 3 | 2 | 1 | 3 | 1 | 5 |
| Cerebellar Lesion | 1 | 3 | 0 | 3 | 1 | 1 | 0 | 3 | 4 |
| Ethmoid Paperboard Fracture | 0 | 4 | 4 | 0 | 0 | 0 | 4 | 0 | 4 |
| Pons Ischemia | 2 | 2 | 0 | 2 | 2 | 0 | 4 | 0 | 4 |
| Cerebral Fissure | 0 | 3 | 0 | 2 | 1 | 0 | 3 | 0 | 3 |
| Demyelination | 2 | 1 | 0 | 0 | 3 | 1 | 2 | 0 | 3 |
| Widened Ventricles | 2 | 1 | 0 | 3 | 0 | 0 | 0 | 3 | 3 |
| Widened Subdural Space | 2 | 1 | 1 | 0 | 2 | 0 | 3 | 0 | 3 |
| Cisterna Magna Cyst | 1 | 2 | 1 | 1 | 1 | 0 | 3 | 0 | 3 |
| Cerebral Hemisphere Compression | 0 | 2 | 1 | 1 | 0 | 0 | 1 | 1 | 2 |
| Bony Process | 2 | 0 | 1 | 0 | 1 | 0 | 1 | 1 | 2 |
| Arterial Thickening | 1 | 1 | 0 | 1 | 1 | 1 | 0 | 1 | 2 |
| Pineal Gland Enlargement | 0 | 2 | 1 | 0 | 1 | 0 | 2 | 0 | 2 |
| Dysplasia | 0 | 1 | 0 | 1 | 0 | 1 | 0 | 0 | 1 |
| Cerebellar Compression | 0 | 1 | 0 | 1 | 0 | 1 | 0 | 0 | 1 |
| Negative Occupancy Effect | 0 | 1 | 0 | 0 | 1 | 0 | 0 | 1 | 1 |
| Arachnoid Granules | 0 | 1 | 0 | 0 | 1 | 0 | 0 | 1 | 1 |
| Widened Tentorium | 1 | 0 | 0 | 0 | 1 | 0 | 1 | 0 | 1 |
| Widened Lambdoid Sutures | 1 | 0 | 0 | 1 | 0 | 0 | 1 | 0 | 1 |
| Ventricular Reduction | 0 | 1 | 0 | 1 | 0 | 0 | 1 | 0 | 1 |
| Adenoid Hypertrophy | 1 | 0 | 1 | 0 | 0 | 0 | 1 | 0 | 1 |
| Cystic Pineal Gland | 1 | 0 | 0 | 1 | 0 | 0 | 0 | 1 | 1 |
| Frontal Lobe Cyst | 1 | 0 | 0 | 1 | 0 | 0 | 1 | 0 | 1 |
| Neuroma | 0 | 1 | 0 | 1 | 0 | 0 | 1 | 0 | 1 |
| Thyroid Cartilage Fracture | 1 | 0 | 0 | 0 | 1 | 0 | 0 | 1 | 1 |
| Capsular Infarction | 0 | 1 | 0 | 1 | 0 | 0 | 1 | 0 | 1 |
| Massive Hemorrhage | 0 | 1 | 0 | 1 | 0 | 0 | 0 | 1 | 1 |

**Supplementary Table 10. Statistics of the disorder-containing scans in the prospective dataset.** The prospective dataset comprises 2,709 scans with 116 types of disorders. These scans were obtained from the PLAGH and span from Jul. 2019 to Aug. 2021.

| Prospective | | | | | | | | |
|---|---|---|---|---|---|---|---|---|
| | **Gender** | | **Age** | | | **Inspection time** | | **Total** |
| | **Male** | **Female** | **~44** | **44~59** | **59~** | **2019~2020** | **2020~** | **-** |
| Ischemia | 261 | 212 | 8 | 55 | 410 | 162 | 311 | 473 |
| Malacia Foci | 307 | 153 | 37 | 83 | 340 | 181 | 279 | 460 |
| Parenchymal Hemorrhage | 146 | 74 | 48 | 45 | 127 | 76 | 144 | 220 |
| Pneumocephaly | 125 | 88 | 67 | 63 | 83 | 107 | 106 | 213 |
| Subarachnoid Hemorrhage | 103 | 110 | 37 | 39 | 137 | 72 | 141 | 213 |
| Brainswelling | 112 | 81 | 77 | 33 | 83 | 61 | 132 | 193 |
| Effusion | 115 | 53 | 29 | 32 | 107 | 65 | 103 | 168 |
| Soft Tissue Swelling | 79 | 63 | 59 | 21 | 62 | 50 | 92 | 142 |
| Subdural Hematoma | 85 | 38 | 21 | 19 | 83 | 40 | 83 | 123 |
| Calcification | 44 | 71 | 25 | 27 | 63 | 43 | 72 | 115 |
| Lacunar Infarction | 73 | 36 | 2 | 31 | 76 | 37 | 72 | 109 |
| Contusion | 78 | 29 | 45 | 21 | 41 | 31 | 76 | 107 |
| Basal Ganglia Cerebral Infarction | 67 | 25 | 5 | 26 | 61 | 29 | 63 | 92 |
| Space Occupying Lesions | 42 | 48 | 29 | 28 | 33 | 20 | 70 | 90 |
| Intraventricular Hemorrhage | 45 | 38 | 21 | 12 | 50 | 22 | 61 | 83 |
| Sclerosis | 52 | 29 | 6 | 8 | 67 | 42 | 39 | 81 |
| Tubercle | 33 | 45 | 30 | 20 | 28 | 33 | 45 | 78 |
| Subcutaneous Hematoma | 38 | 40 | 18 | 4 | 56 | 27 | 51 | 78 |
| Midline Shift | 52 | 22 | 10 | 34 | 30 | 33 | 41 | 74 |
| Cavum Septum Pellucidum | 43 | 29 | 22 | 19 | 31 | 24 | 48 | 72 |
| Cerebral Infarction of Caudate Nucleus Head | 43 | 29 | 4 | 22 | 46 | 22 | 50 | 72 |
| Arachnoid Cyst | 54 | 13 | 39 | 13 | 15 | 16 | 51 | 67 |
| Cisterna Magna | 59 | 7 | 26 | 9 | 31 | 14 | 52 | 66 |
| Artifacts Shadow | 40 | 25 | 13 | 10 | 42 | 14 | 51 | 65 |
| Brain Atrophy | 43 | 20 | 1 | 7 | 55 | 7 | 56 | 63 |
| Epidural Hematoma | 37 | 25 | 24 | 17 | 21 | 21 | 41 | 62 |
| Basal Ganglia Ischemia | 35 | 26 | 8 | 18 | 35 | 19 | 42 | 61 |
| Minor Hemorrhage | 38 | 22 | 14 | 14 | 32 | 19 | 41 | 60 |
| Multiple Cerebral Infarctions | 39 | 20 | 1 | 14 | 44 | 25 | 34 | 59 |
| Cerebral Edema | 33 | 25 | 7 | 16 | 35 | 17 | 41 | 58 |
| Ventriculomegaly | 32 | 23 | 13 | 11 | 31 | 26 | 29 | 55 |
| Paraventricular Ischemia | 35 | 18 | 3 | 9 | 41 | 11 | 42 | 53 |
| Atherosclerosis | 30 | 18 | 0 | 3 | 45 | 26 | 22 | 48 |
| Scalp Hematoma | 18 | 30 | 23 | 9 | 16 | 9 | 39 | 48 |
| Periventricular Cerebral Infarction | 39 | 8 | 1 | 19 | 27 | 14 | 33 | 47 |
| Subdural Hemorrhage | 23 | 24 | 12 | 7 | 28 | 22 | 25 | 47 |
| Senile Encephalopathy | 35 | 9 | 1 | 1 | 42 | 3 | 41 | 44 |
| Hydrocephalus | 30 | 14 | 17 | 10 | 17 | 12 | 32 | 44 |
| Occipital Fracture | 26 | 16 | 23 | 6 | 13 | 21 | 21 | 42 |
| Osteoma | 13 | 28 | 5 | 6 | 30 | 12 | 29 | 41 |

| Prospective | | | | | | | | |
|---|---|---|---|---|---|---|---|---|
| | Gender | | Age | | | Inspection time | | Total |
| | Male | Female | ~44 | 44~59 | 59~ | 2019~2020 | 2020~ | - |
| Parietal Fracture | 27 | 8 | 24 | 6 | 5 | 10 | 25 | 35 |
| Frontal Lobe Ischemia | 17 | 18 | 1 | 8 | 26 | 15 | 20 | 35 |
| Parenchymal Hematoma | 13 | 22 | 10 | 4 | 21 | 7 | 28 | 35 |
| Herniation | 21 | 11 | 3 | 12 | 17 | 7 | 25 | 32 |
| Lipoma | 11 | 19 | 5 | 4 | 21 | 10 | 20 | 30 |
| Arteriosclerosis | 17 | 6 | 1 | 2 | 20 | 14 | 9 | 23 |
| Frontal Bone Fracture | 18 | 5 | 17 | 5 | 1 | 8 | 15 | 23 |
| Hemispheric Infarction | 16 | 7 | 0 | 15 | 8 | 3 | 20 | 23 |
| Absence Of Bone | 9 | 13 | 5 | 6 | 11 | 9 | 13 | 22 |
| Temporal Lobe Cerebral Infarction | 12 | 9 | 2 | 5 | 14 | 7 | 14 | 21 |
| Frontal Lobe Cerebral Infarction | 11 | 10 | 0 | 6 | 15 | 2 | 19 | 21 |
| Corona Radiata Cerebral Infarction | 14 | 4 | 0 | 3 | 15 | 12 | 6 | 18 |
| Parietal Lobe Ischemia | 10 | 7 | 0 | 5 | 12 | 4 | 13 | 17 |
| Occipital Lobe Cerebral Infarction | 16 | 0 | 0 | 2 | 14 | 4 | 12 | 16 |
| Zygomatic Fracture | 7 | 8 | 0 | 3 | 12 | 5 | 10 | 15 |
| Temporal Bone Fracture | 13 | 2 | 12 | 1 | 2 | 4 | 11 | 15 |
| Parietal Lobe Cerebral Infarction | 10 | 5 | 2 | 1 | 12 | 2 | 13 | 15 |
| Malformation | 4 | 10 | 3 | 2 | 9 | 1 | 13 | 14 |
| Widened Cisterna Magna | 6 | 8 | 7 | 4 | 3 | 2 | 12 | 14 |
| Cystic Foci | 6 | 8 | 3 | 7 | 4 | 7 | 7 | 14 |
| Wallerian Degeneration | 11 | 2 | 2 | 2 | 9 | 2 | 11 | 13 |
| Empty Sella | 5 | 8 | 1 | 0 | 12 | 4 | 9 | 13 |
| Septum Pellucidum Cyst | 5 | 8 | 7 | 4 | 2 | 2 | 11 | 13 |
| Skull Fracture | 8 | 5 | 8 | 2 | 3 | 0 | 13 | 13 |
| Metal Shadow | 3 | 9 | 2 | 4 | 6 | 2 | 10 | 12 |
| Zygomatic Arch Fracture | 4 | 8 | 0 | 3 | 9 | 6 | 6 | 12 |
| Centrum Semiovale Ischemia | 3 | 9 | 0 | 6 | 6 | 3 | 9 | 12 |
| Brainstem Ischemia | 8 | 4 | 1 | 1 | 10 | 5 | 7 | 12 |
| Osseous Defects | 6 | 5 | 3 | 0 | 8 | 5 | 6 | 11 |
| Widened Ventricles | 7 | 3 | 2 | 1 | 7 | 4 | 6 | 10 |
| Corona Radiata Ischemia | 7 | 3 | 1 | 2 | 7 | 2 | 8 | 10 |
| Cerebral Fissure | 7 | 1 | 2 | 3 | 3 | 2 | 6 | 8 |
| Fat Density | 2 | 6 | 1 | 0 | 7 | 4 | 4 | 8 |
| Insular Infarction | 4 | 4 | 0 | 0 | 8 | 4 | 4 | 8 |
| Cerebral White Matter Degeneration | 3 | 4 | 0 | 0 | 7 | 6 | 1 | 7 |
| Metastases | 1 | 6 | 0 | 2 | 5 | 4 | 3 | 7 |
| Thalamic Cerebral Infarction | 5 | 2 | 0 | 4 | 3 | 2 | 5 | 7 |
| Old Cerebral Infarction | 4 | 3 | 0 | 4 | 3 | 1 | 6 | 7 |
| Widened Subdural Space | 3 | 3 | 0 | 1 | 5 | 2 | 4 | 6 |
| Meningioma | 2 | 4 | 0 | 4 | 2 | 1 | 5 | 6 |

| Prospective | | | | | | | | |
|---|---|---|---|---|---|---|---|---|
| | Gender | | Age | | | Inspection time | | Total |
| | **Male** | **Female** | **~44** | **44~59** | **59~** | **2019~2020** | **2020~** | **-** |
| Centrum Semiovale Cerebral Infarction | 4 | 2 | 0 | 1 | 5 | 2 | 4 | 6 |
| Bony Process | 0 | 5 | 0 | 5 | 0 | 3 | 2 | 5 |
| Fat Deposition | 2 | 3 | 0 | 0 | 5 | 1 | 4 | 5 |
| Tumour | 3 | 2 | 1 | 1 | 3 | 2 | 3 | 5 |
| Multiple Hemorrhages | 3 | 2 | 0 | 1 | 4 | 3 | 2 | 5 |
| Enhancement Foci | 1 | 3 | 0 | 0 | 4 | 2 | 2 | 4 |
| Widened Tentorium | 1 | 3 | 0 | 1 | 3 | 0 | 4 | 4 |
| Cisterna Magna Cyst | 4 | 0 | 0 | 0 | 4 | 2 | 2 | 4 |
| Cerebellar Hemisphere Infarction | 3 | 1 | 0 | 0 | 4 | 2 | 2 | 4 |
| Arterial Thickening | 2 | 1 | 0 | 0 | 3 | 0 | 3 | 3 |
| Artery Tortuosity | 3 | 0 | 0 | 1 | 2 | 0 | 3 | 3 |
| Widened Arterial | 3 | 0 | 0 | 1 | 2 | 1 | 2 | 3 |
| Sphenoid Fracture | 3 | 0 | 0 | 3 | 0 | 0 | 3 | 3 |
| Brainstem Infarction | 3 | 0 | 0 | 0 | 3 | 2 | 1 | 3 |
| Subacute Cerebral Infarction | 3 | 0 | 0 | 1 | 2 | 1 | 2 | 3 |
| Epidural Hemorrhage | 3 | 0 | 2 | 0 | 1 | 0 | 3 | 3 |
| Brainstem Swelling | 2 | 0 | 0 | 2 | 0 | 0 | 2 | 2 |
| Arterial Cilia | 0 | 2 | 0 | 0 | 2 | 1 | 1 | 2 |
| Widened Subarachnoid Space | 0 | 2 | 0 | 0 | 2 | 1 | 1 | 2 |
| Myeloma | 2 | 0 | 0 | 0 | 2 | 1 | 1 | 2 |
| Dysplasia | 0 | 1 | 0 | 0 | 1 | 0 | 1 | 1 |
| Cerebral Hemisphere Compression | 1 | 0 | 0 | 0 | 1 | 1 | 0 | 1 |
| Arachnoid Granules | 0 | 1 | 0 | 0 | 1 | 0 | 1 | 1 |
| Leukoaraiosis | 0 | 1 | 0 | 0 | 1 | 0 | 1 | 1 |
| Cerebral Cortical Atrophy | 1 | 0 | 0 | 0 | 1 | 0 | 1 | 1 |
| Ventricular Reduction | 0 | 1 | 0 | 0 | 1 | 0 | 1 | 1 |
| Pineal Gland Enlargement | 1 | 0 | 0 | 0 | 1 | 0 | 1 | 1 |
| Sella Turcica Enlargement | 0 | 1 | 0 | 0 | 1 | 0 | 1 | 1 |
| Choroid Cyst | 0 | 1 | 0 | 0 | 1 | 1 | 0 | 1 |
| Frontal Lobe Cyst | 1 | 0 | 0 | 0 | 1 | 0 | 1 | 1 |
| Facial Bone Fracture | 1 | 0 | 1 | 0 | 0 | 0 | 1 | 1 |
| Pons Ischemia | 1 | 0 | 0 | 0 | 1 | 0 | 1 | 1 |
| Insula Ischemia | 0 | 1 | 0 | 0 | 1 | 1 | 0 | 1 |
| Minor Haematoma | 0 | 1 | 0 | 1 | 0 | 0 | 1 | 1 |
| Massive Hemorrhage | 1 | 0 | 0 | 0 | 1 | 0 | 1 | 1 |
| Cerebellar Vermis Hemorrhage | 1 | 0 | 0 | 1 | 0 | 1 | 0 | 1 |

**Supplementary Table 11. Statistics of the disorder-containing scans in the cross-center dataset.** The cross-center dataset comprises 327 scans with 46 types of disorders. These scans were obtained from the PLAGH and span from Apr. 2018 to May. 2019.

| Prospective | | | | | | | | |
|---|---|---|---|---|---|---|---|---|
| | Gender | | Age | | | Inspection time | | Total |
| | Male | Female | ~44 | 44~59 | 59~ | 2018~2019 | 2019~ | - |
| Lacunar Infarction | 67 | 33 | 0 | 8 | 92 | 66 | 34 | 100 |
| Senile Encephalopathy | 65 | 34 | 0 | 6 | 93 | 66 | 33 | 99 |
| Subarachnoid Hemorrhage | 61 | 35 | 13 | 31 | 52 | 62 | 34 | 96 |
| Parenchymal Hemorrhage | 60 | 28 | 11 | 25 | 52 | 57 | 31 | 88 |
| Contusion | 49 | 15 | 13 | 26 | 25 | 47 | 17 | 64 |
| Malacia Foci | 40 | 16 | 3 | 5 | 48 | 41 | 15 | 56 |
| Subdural Hematoma | 45 | 7 | 8 | 20 | 24 | 34 | 18 | 52 |
| Parenchymal Hematoma | 32 | 11 | 10 | 11 | 22 | 18 | 25 | 43 |
| Intraventricular Hemorrhage | 26 | 15 | 4 | 12 | 25 | 24 | 17 | 41 |
| Soft Tissue Injury | 30 | 9 | 10 | 14 | 15 | 20 | 19 | 39 |
| Epidural Hematoma | 20 | 2 | 11 | 8 | 3 | 15 | 7 | 22 |
| Leukoaraiosis | 12 | 9 | 0 | 1 | 20 | 11 | 10 | 21 |
| Brain Atrophy | 12 | 7 | 0 | 0 | 19 | 8 | 11 | 19 |
| Pneumocephaly | 14 | 5 | 6 | 10 | 3 | 14 | 5 | 19 |
| Effusion | 11 | 6 | 3 | 3 | 11 | 9 | 8 | 17 |
| Scalp Hematoma | 4 | 10 | 1 | 2 | 11 | 10 | 4 | 14 |
| Brainswelling | 8 | 4 | 3 | 4 | 5 | 6 | 6 | 12 |
| Herniation | 6 | 3 | 3 | 0 | 6 | 4 | 5 | 9 |
| Soft Tissue Swelling | 7 | 0 | 2 | 3 | 2 | 6 | 1 | 7 |
| Occipital Lobe Cerebral Infarction | 3 | 3 | 0 | 1 | 5 | 3 | 3 | 6 |
| Basal Ganglia Cerebral Infarction | 2 | 3 | 0 | 2 | 3 | 1 | 4 | 5 |
| Arachnoid Cyst | 4 | 0 | 2 | 1 | 1 | 3 | 1 | 4 |
| Frontal Lobe Cerebral Infarction | 2 | 2 | 0 | 1 | 3 | 0 | 4 | 4 |

| Prospective | | | | | | | | |
|---|---|---|---|---|---|---|---|---|
| | **Gender** | | **Age** | | | **Inspection time** | | **Total** |
| | **Male** | **Female** | **~44** | **44~59** | **59~** | **2018~2019** | **2019~** | **-** |
| Calcification | 2 | 1 | 0 | 1 | 2 | 1 | 2 | 3 |
| Parietal Lobe Cerebral Infarction | 0 | 3 | 0 | 1 | 2 | 0 | 3 | 3 |
| Cerebellar Hemisphere Infarction | 3 | 0 | 0 | 1 | 2 | 3 | 0 | 3 |
| Temporal Lobe Cerebral Infarction | 2 | 1 | 0 | 0 | 3 | 0 | 3 | 3 |
| Wallerian Degeneration | 1 | 2 | 0 | 0 | 3 | 3 | 0 | 3 |
| Insular Infarction | 1 | 2 | 0 | 1 | 2 | 0 | 3 | 3 |
| Hemispheric Infarction | 2 | 0 | 0 | 0 | 2 | 1 | 1 | 2 |
| Cerebral Edema | 2 | 0 | 0 | 0 | 2 | 2 | 0 | 2 |
| Subdural Hemorrhage | 1 | 0 | 0 | 0 | 1 | 1 | 0 | 1 |
| Multiple Hemorrhages | 1 | 0 | 0 | 1 | 0 | 1 | 0 | 1 |
| Subcutaneous Hematoma | 1 | 0 | 0 | 0 | 1 | 0 | 1 | 1 |
| Minor Hemorrhage | 1 | 0 | 1 | 0 | 0 | 1 | 0 | 1 |
| Brainstem Infarction | 1 | 0 | 0 | 1 | 0 | 1 | 0 | 1 |
| Temporal Bone Fracture | 1 | 0 | 1 | 0 | 0 | 0 | 1 | 1 |
| Fat Density | 0 | 1 | 0 | 0 | 1 | 1 | 0 | 1 |
| Midline Shift | 0 | 1 | 0 | 0 | 1 | 1 | 0 | 1 |
| Thalamic Cerebral Infarction | 1 | 0 | 0 | 1 | 0 | 1 | 0 | 1 |
| Cerebellar Vermis Hemorrhage | 1 | 0 | 1 | 0 | 0 | 1 | 0 | 1 |
| Subacute Cerebral Infarction | 1 | 0 | 0 | 0 | 1 | 1 | 0 | 1 |
| Malformation | 0 | 1 | 1 | 0 | 0 | 0 | 1 | 1 |
| Hydrocephalus | 1 | 0 | 0 | 0 | 1 | 0 | 1 | 1 |
| Centrum Semiovale Cerebral Infarction | 1 | 0 | 0 | 0 | 1 | 0 | 1 | 1 |
| Periventricular Cerebral Infarction | 1 | 0 | 0 | 0 | 1 | 0 | 1 | 1 |

**Supplementary Table 12. Performance of baseline methods on the pulmonary CT test dataset for pulmonary disorder detection.** The results of each method were obtained by averaging their performance on different types of pulmonary disorders.

| Pulmonary CT | | | |
|---|---|---|---|
| | **AUC** | **Sensitivity** | **Specificity** |
| Auto-Encoder | 0.719 (0.716, 0.722) | 0.547 (0.541, 0.552) | 0.832 (0.830, 0.833) |
| AnoGAN | 0.819 (0.818, 0.823) | 0.680 (0.674, 0.685) | 0.837 (0.835, 0.839) |
| GANomaly | 0.846 (0.845, 0.849) | 0.717 (0.712, 0.723) | 0.839 (0.836, 0.841) |
| pix2pix | 0.746 (0.744, 0.750) | 0.562 (0.556, 0.568) | 0.831 (0.830, 0.833) |
| Cycle-GAN | 0.828 (0.827, 0.832) | 0.686 (0.680, 0.692) | 0.837 (0.835, 0.839) |
| Ours | 0.893 (0.891, 0.894) | 0.838 (0.834, 0.842) | 0.854 (0.853, 0.855) |

**Supplementary Table 13. Comparison of model performance with different encoder-decoder architectures.** The comparison experiment was conducted on the cross-center test datasets.

| Architecture | AUC |
|---|---|
| Ours (8 Resnet Blocks) | 0.866 |
| 6 Resnet Blocks | 0.852 |
| 9 Resnet Blocks | 0.868 |
| Unet | 0.841 |

**Supplementary Table 14. comparison of model performance with perceptual loss.** This table presents an analysis of the performance metrics when perceptual loss is calculated from different layers of VGG-19 and ResNet34 networks. VGG-19, perceptual loss was computed using the output from the first ReLU activation layer in each of the five blocks, while for ResNet34, it was derived from the output of the last layer in each of the four blocks. The comparison experiment was conducted on the cross-center test datasets.

| Architecture | AUC |
|---|---|
| VGG-19 | 0.866 |
| ResNet-34 | 0.865 |

**Supplementary Table 15. Network architecture used for the de-disorder network.** For each output of the convolution layer, we applied an instance norm layer and a ReLU nonlinear activation layer for processing. The details of the Residual block are shown in Supplementary Fig. 2.

| Layer | Activation size |
|---|---|
| Input | 6×512×512 |
| 64×7×7 conv, stride 1 | 64×512×512 |
| 128×4×4 conv, stride 2 | 128×256×256 |
| 256×4×4 conv, stride 2 | 256×128×128 |
| (Residual block, 256 filters) × 8 | 256×128×128 |
| 128×4×4 conv, stride 1/2 | 128×256×256 |
| 64×3×3 conv, stride 1/2 | 64×512×512 |
| 3×7×7 conv, stride 1 | 3×512×512 |

**Supplementary Table 16. The parameter settings of our system.**

| | **Parameters** | **Description** | **Value** |
|---|---|---|---|
| **Input related** | input size | The size of one CT slice to be fed into the networks | $512 \times 512$ |
| | WW | The value of window width | 80 |
| | WL | The value of window location | 40 |
| | backbone | The backbone network of the DeDN | Shown in Supplementary Table 15 |
| | $K$ | The image fed into DeDN is divided into $K \times K$ grids | 4 |
| | $n_e$ | the number of image edge maps | 3 |
| **DeDN related** | $\lambda_{\ell_1}$ | The hyper-parameter in Eq. (3) | 1 |
| | $\lambda_{adv}$ | The hyper-parameter in Eq. (3) | 0.5 |
| | $\lambda_p$ | The hyper-parameter in Eq. (3) | 0.1 |
| | $\lambda_s$ | The hyper-parameter in Eq. (3) | 10 |
| **DRN related** | backbone | The backbone network of the DRN | ResNet-50 |
| | $\lambda_c$ | The hyper-parameter in Eq. (9) | 1 |
| | $\lambda_d$ | The hyper-parameter in Eq. (9) | 0.5 |
| **Optimization related** | optimizer | The iterative method for optimizing the objective function | SGD |
| | lr | Learning rate to optimize network | 0.01 |
| | init lr | The initial learning rate to warm up the network | 0.003 |
| | batch size | The number of CT slices utilized in one training iteration | 40 |
| | momentum factor | Momentum factor for accelerating gradients vectors | 0.9 |
| | L2 penalty | The regularization ratio to prevent overfitting | 5e-4 |
| **Visualization related** | $t$ | The normal pixel bias range on a difference image | 10 |
| | $s$ | The edge length of the region used to smooth one pixel | 3 |

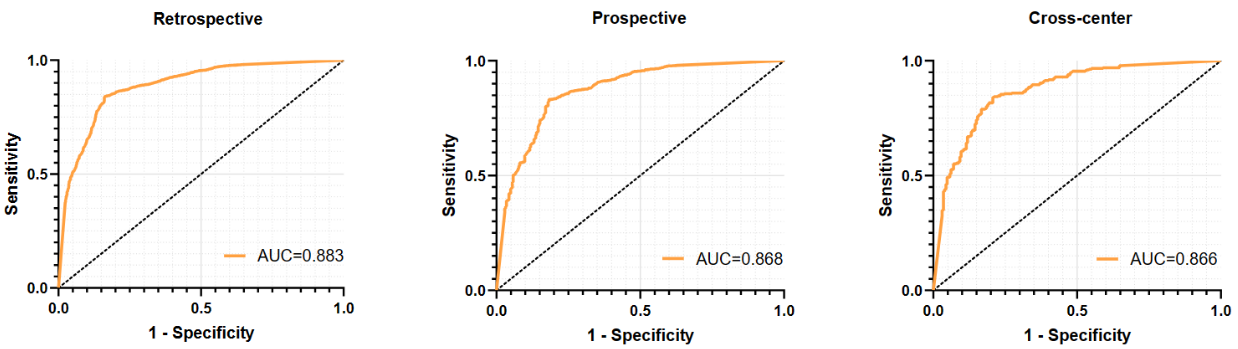

**Supplementary Figure 1. ROC curves for the disorder recognition on the retrospective, prospective and cross-center test datasets.**

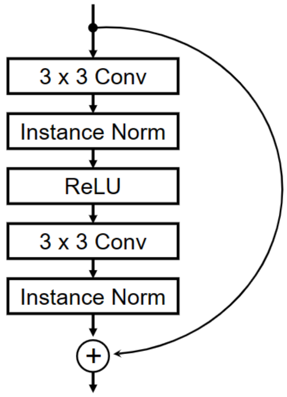

**Supplementary Figure 2. Residual block design used in the de-disorder network.**
